# *MARK2* variants cause autism spectrum disorder *via* the downregulation of WNT/β-catenin signaling pathway

**DOI:** 10.1101/2024.04.24.24304501

**Authors:** Maolei Gong, Jiayi Li, Yijun Liu, Matheus Vernet Machado Bressan Wilke, Qian Li, Haoran Liu, Chen Liang, Joel A Morales-Rosado, Ana S.A. Cohen, Susan S. Hughes, Bonnie R. Sullivan, Valerie Waddell, Marie-José H. van den Boogaard, Richard H. van Jaarsveld, Ellen van Binsbergen, Koen L van Gassen, Tianyun Wang, Susan M. Hiatt, Michelle D. Amaral, Whitley V. Kelley, Jianbo Zhao, Weixing Feng, Changhong Ren, Yazhen Yu, Nicole J Boczek, Matthew J. Ferber, Carrie Lahner, Sherr Elliott, Yiyan Ruan, Mignot Cyril, Boris Keren, Hua Xie, Xiaoyan Wang, Bernt Popp, Christiane Zweier, Juliette Piard, Christine Coubes, Frederic Tran Mau-Them, Hana Safraou, Micheil Innes, Julie Gauthier, Jacques Michaud, Daniel C. Koboldt, Odent Sylvie, Marjolaine Willems, Wen-Hann Tan, Benjamin Cogne, Claudine Rieubland, Dominique Braun, Scott Douglas McLean, Konrad Platzer, Pia Zacher, Henry Oppermann, Lucie Evenepoel, Pierre Blanc, Laïla El Khattabi, Neshatul Haque, Nikita R. Dsouza, Michael T. Zimmermann, Raul Urrutia, Eric W Klee, Yiping Shen, Hongzhen Du, Zailong Qin, Chang-Mei Liu, Xiaoli Chen

## Abstract

*MARK2*, a member of the evolutionarily conserved PAR1/MARK serine/threonine kinase family, has been identified as a novel risk gene for autism spectrum disorder (ASD) based on the enrichment of *de novo* loss-of-function (Lof) variants in large-scale sequencing studies of ASD individuals. However, the features shared by affected individuals and the molecular mechanism of *MARK2* variants during early neural development remained unclear. Here, we report 31 individuals carrying heterozygous *MARK2* variants and presenting with ASD, other neurodevelopmental disorders, and typical facial dysmorphisms. Lof variants predominate (81%) in affected individuals, while computational analysis and *in vitro* transfection assay also point to *MARK2* loss resulting from missense variants. Using patient-derived and CRISPR-engineered isogenic induced pluripotent stem cells (iPSCs), and *Mark*2^+/-^ (HET) mice, we show that *MARK2* loss leads to systemic neurodevelopmental deficits, including anomalous polarity in neural rosettes, imbalanced proliferation and differentiation in neural progenitor cells (NPCs), abnormal cortical development and ASD-like behaviors in mice. Further using RNA-Seq and lithium treatment, we link *MARK2* loss to the downregulated WNT/β-catenin signaling pathway and identify lithium as a potential drug for treating *MARK2*-related ASD.

## Introduction

Autism spectrum disorder (ASD) is one of neurodevelopmental disorders (NDDs) marked by difficulties in social communication and repetitive behaviors,^1^ and affects 0.7-1.69% of children worldwide.^2, 3^ ASD shows high heritability and 5-30% of affected individuals have a Mendelian disorder.^4, 5^ In light of advances in genomic technologies, over 100 high-confidence ASD genes have been identified by large-scale sequencing studies of autism cohorts.^6–8^ Most of these causative genes are highly intolerant to loss-of-function (Lof) variants in the human genome, but are enriched for *de novo* variants in ASD individuals.^6–8^

Microtubule affinity-regulating kinase 2 (*MARK2*, OMIM600526), also known as partitioning-defective 1b (PAR1b), is a member of the evolutionarily conserved PAR1/MARK serine/threonine kinase family,^9, 10^ and it plays essential roles in the central nervous system (CNS) via several cell biological functions.^11–16^ It has been shown in primary hippocampal neurons that *MARK2* negatively modulates neuronal polarity and dendritic development, and neurons with *MARK2* depletion exhibit unpolarized dendritic overgrowth with multiplex rather than single axons.^11, 12^ Additionally, downregulation of *MARK2* by *in utero* electroporation inhibits neuronal migration in the mouse cortex, which shows that unpolarized neurons accumulate in the intermediate zone (IZ) of the developing cerebral cortex.^13, 14^ Recently, Zhou et al. performed an integrated analysis of the largest set of sequencing data from 42,607 ASD patients and identified *MARK2* as a novel moderate-risk gene for ASD based on the enrichment of rare *de novo* and inherited Lof variants in affected patients.^17^ However, the phenotypic and variant spectrum of *MARK2*-related ASD, as well as the underlying pathogenic mechanisms of *MARK2* variants during early neural development, remain unclear.

Herein, we describe a cohort of 31 individuals carrying clinically relevant *MARK2* variants, mostly Lof variants (25/31), which were gathered through GeneMatcher.^18^ All affected individuals presented with distinct facial dysmorphisms, ASD and other NDD phenotypes. Using patient-derived induced pluripotent stem cells (iPSCs) combining with CRISPR-engineered isogenic iPSCs, we revealed the cellular phenotypes and transcriptomic profiles of mutant iPSC-derived neural progenitor cells (NPCs). Furthermore, using *Mark2*^+/-^ (HET) mice, we recapitulated the pathophysiological deficits observed in mutant iPSC-derived NPCs and the ASD-related behaviors presented by affected individuals. Importantly, we revealed the downregulation of WNT/β-catenin signaling pathway as the molecular mechanism of *MARK2* variant, which could be rescued by lithium at both cellular and tissue levels indicating a potential therapeutic approach for *MARK2*-related ASD.

## Materials and methods

### Ethics statement

The study was approved by the ethics committees of the respective institutions, including the Capital Institute of Pediatrics (SHERLL202001), Mayo Clinic (IRB#19-003389) and Maternal and Child Health Hospital of Guangxi Zhuang Autonomous Region (2017-3-11). This international individual cohort carrying *MARK2* variant were recruited by GeneMatcher data sharing platform.^18^ Informed consent was obtained from the parents of some individuals to allow their clinical information, genotypic data and facial photos to be published. The detailed phenotypes were recorded by the referring clinician. Sample collection and patient consent were in accordance with the tenets of the Declaration of Helsinki.

### Detection of *MARK2* variants

*MARK2* variants were detected in peripheral blood samples by proband-only whole exome sequencing (WES) or Trio-WES, whole-genome sequencing (WGS) combined with Sanger sequencing. WES was performed with commercial exome capture kits and was followed by sequencing on the Illumina NovaSeq 6000 platform.

Variants were annotated based on transcript NM_001039469.35 using the reference human genome GRCh37(hg19) version. The inheritance of *MARK2* variants was confirmed in all families by Sanger sequencing. Two pairs of primers were designed to exclude amplification bias during PCR. For missense variants, only those that were predicted as deleterious variants by more than two software programs were included. The pathogenicity of each candidate mutation was classified according to the 2015 guidelines of American College of Medical Genetics and Genomics (ACMG).^19^ The identities of biological parents were confirmed for all affected individuals before using the PS2 criteria. The criteria for PP3^20^ and PVS1^21^ evidences were modified based on the new ACMG guidelines.

### Expression vector construction, cell culture, transient transfection

Two splicing variants (c.337+1G>T, c.1514+2T>G) and six missense variants (A80V, G135R, F194S, R302Q, V752A, R764P) were chosen for *in vitro* transient transfection and functional assay. For splicing variant, two genomic fragments covering wild-type (WT) *MARK2* c.337+1G allele (1039bp; containing 1bp of intron 2, 54bp of exon 3, 541bp of partial intron 3, 49bp of exon 4, 327bp of intron 4 and 66bp of exon 5, and 1bp of intron 5; see **Supplementary Table 1** for primers) or covering the c.1514+2T allele (2441bp; containing 79bp of exon 13, 278bp of intron 13, 98bp of exon 14, 827bp of intron 14, 162bp of exon 639bp of intron 16 and 258bp of exon 16; see **Supplementary Table 1** for primers) were amplified by standard PCR or overlap-extension PCR and then cloned into the pcDNA3.1 vector. For missense variants, full length WT *MARK2* cDNA (2367bp, NM_001039469.2) was synthesized and cloned into pCMV-EGFP-Neo vector using two primers containing KpnI and Agel restriction enzyme sites. The mutant vectors were constructed by site-specific mutant primers and overlap-extension PCR (**Supplementary Table 1**). All vectors were verified by sequencing.

Human embryonic kidney (HEK293T) cells and HeLa cells (ATCC, Manassas, VA) were grown in minimum Eagle’s medium supplemented with 10% fetal bovine serum and antibiotics. The cells were grown to 70%–90% confluence before transient transfection (Lipofectamine 2000 reagent, Invitrogen, CA, USA). 48h later, transfected cells were washed twice with cool 1x PBS, and then collected for RNA extraction (splicing variants), protein lysis and western blotting (missense variants). Total RNA was extracted (TRIzol, Omega, GA, USA) and reverse transcribed into cDNA (SuperScript reverse transcription kit, 18064071, Thermo, MA, USA) following with standard PCR **(Supplementary Table 1 for primers**). The PCR products were visualized on a 2% agarose gel and then purified for Sanger sequencing.

### Model construction and calculation

The canonical transcript, NM_001039469.2 was used. The structure of the kinase-associated domain 1 (KA1) of human MARK2 is unknown, but the experimental structures of all human KA1 domains are sufficiently identical (71%) to generate a high-confidence KA1 model through homology-based methods.^22^ The structures of the kinase and UBA domains of MARK2 have been experimentally determined (PDB ID: 1zmu).^23^ These independently generated models were integrated using information from AlphaFold2.^24^

Regions of the protein that do not adopt secondary structures and are likely to be intrinsically disordered regions (IDRs) were loop modeled using Modeller v10.2.^25^ The combined 3D model was used for visualization and structure-based calculations of stability alterations due to genomic variants.^26^ We assessed short linear motifs (SLiMs) of each missense variant by programmatically accessing the web API of ELM.^27^ In addition, we calculated the local stability (ΔΔGfold) and the balance of favorable and unfavorable local contacts, termed frustration for missense variants.

### Generation of iPSC lines from two affected individuals

Peripheral blood mononuclear cells (PBMCs) were reprogrammed into iPSCs using Invitrogen a CytoTune-iPS 2.0 Sendai Reprogramming Kit (Invitrogen) as we previously reported.^28^ In brief, PBMCs were seeded in 12-well plates and cultured for 14 days in PBMC medium. Then, 2.5×10^5^ PBMCs were infected with Sendai virus and cultured for 48 h at 37°C. Then, the infected cells were added to mouse embryonic fibroblast (MEF) feeders in iPSC medium. After 2 weeks, individual iPSC colonies were picked manually, transferred to Matrigel (Corning)-coated dishes, and maintained in mTeSR medium (Stem Cell Technologies). Routine testing revealed that the cells were mycoplasma negative. After more than 10 passage, the characteristics of the iPSCs, including their pluripotency and genomic background, were analyzed. For pluripotency analysis, we performed AP staining and immunofluorescence staining and assessed teratoma formation. To evaluate genomic background, we performed karyotyping, short tandem repeat (STR) site analysis and array CGH to confirm that the genomes were identical. The relationship between two families was confirmed by STR. Two iPSC lines from two normal unrelated adults we previous reported were used as control iPSCs.^28^ Meanwhile, two isogenic iPSCs were generated using CRISPR/Cas9 gene editing engineering to delete the KA1 domain of *MARK2* (CRISPR-del1 and CRISPR-del2, see **Supplementary Table 1** for gRNA primers).

### Neural differentiation of iPSCs

iPSCs were maintained on Matrigel (BD Biosciences) in mTeSR1 medium (StemCell Technologies) and performed. Between passages 12 and 30, iPSCs were used for neural differentiation as previously described with minor modifications.^29^ In brief, 30, 000 iPSCs cells were used for embryoid bodies (EBs) culture, and after 24h EBs were plant to poly-L-ornithine/laminin-coated plates. And then, EBs were grown in 1:1 mixture of N2- and B27-containing media (N1 medium) supplemented with EGF2 (Gibco, 1mg/mL), Noggin (BD Biosciences, 250ng/mL), SB431542 (selleck, 10mM) and Laminin (Gibco, 0.5mg/mL) for 10 days. For neural differentiation, N1 medium was changed to a 1:1 mixture of N2- and B27-containing media (N2 medium) supplemented with FGF1 (Gibco, 1mg/mL), EGF2 (Gibco, 1 mg/mL), BDNF (Gibco,100μg/mL) and Laminin (Gibco, 0.5mg/mL) for another 3 days. For neurospheres, EBs were grown in suspension culture in DMEM/F12 supplemented with 1× N2 (Gibco), 1×nonessential amino acids (Gibco), Knockout™ SR (Gibco) and Mercaptoethanol (Sigma, 25nM) for 5 days.

### *In vitro* and *in vivo* rescue assay of LiCI and recombinant human WNT3A (rhWNT3A)

For the *in vitro* rescue assay, either 0.7 mM LiCl (sigma) or 100mg/ml rhWNT3A (R&D Systems) were supplemented in the culture media from the first day of differentiation process. The rescue efficacy was evaluated at specific time points through WB and IF.

For the *in vivo* rescue assay, 6-8 month aged female WT mice were allowed to mate freely with male HET mice. Once the vaginal plug was observed at the following morning, the drinking water was replaced with clean drinking water containing LiCI (4.5 mg/L). The brains of fetal mice were collected at E18.5 for IF.

### RNA-Seq

Total RNA was extracted from iPSC-derived NPCs with TRIzol reagent (Invitrogen, 15596018) according to the manufacturer’s protocol. Two micrograms of RNA were used for sequencing library generation using the NEBNext® Ultra™ RNA Library Prep Kit (#E7530L, NEB, USA). The cDNA library was preliminarily quantified using a Qubit® RNA Assay Kit (Qubit® 3.0), and then diluted to 1 ng/μl. Insert size was assessed using the Agilent Bioanalyzer 2100 system (Agilent Technologies, CA, USA), and quantified using StepOnePlus™ RT-PCR. The cDNA library was sequenced with the Illumina HiSeq 2500 platform (Annoroad Gene Tech. Co., Ltd. Beijing) to generate 150bp paired-end sequence reads Read counts and FPKM for genes was extracted using the standard RNA-Seq analysis flowchart. The raw sequence data reported in this paper have been deposited in the Genome Sequence Archive (Genomics, Proteomics & Bioinformatics 2021) in the National Genomics Data Center,^30^ China National Center for Bioinformation/Beijing Institute of Genomics, Chinese Academy of Sciences and is publicly accessible at https://ngdc.cncb.ac.cn/gsa-human (GSA-Human: HRA003632).

### RT-quantitative PCR (qPCR) validation

Total RNA from iPSC-derived NPCs was reverse transcribed into cDNA using the Transcript One-Step gDNA Removal and cDNA Synthesis Kit (TransGen Biotech, Beijing, China), and the diluted (1:100) cDNA was used as a template for qPCR. qPCR was performed on a 7500 FAST Real-Time PCR system (Life Technologies) using a SYBR® Select Master Mix Kit (Life Technologies) with specific primers. Relative gene expression was analyzed by the 2^DΔΔCT^ method, and GAPDH or actin was used as an endogenous control. The RT-qPCR primers are listed in **Supplementary Table 1**.

### Western blot analysis and immunofluorescence staining

Total protein was extracted from cells or tissue using RIPA buffer containing 10mM PMSF (Beyotime Biotechnology, ST505), and the protein concentration was determined using a BCA protein assay kit (Beyotime Biotechnology, P0012). The membranes were blocked in 5% milk in TBST (0.1% Tween 20) and incubated with primary antibodies, at 4°C overnight. The membranes were then washed in TBST for 10 min, 3 times and incubated with secondary antibodies at room temperature for 2 h. An ECL system (Pierce) and Tanon-5200 Chemiluminescent Imaging System (Tanon, China, Shanghai) were used for signal detection.

Brain of E18.5 mouse were postfixed with 4% paraformaldehyde (PFA) overnight and then dehydrated with 30% sucrose. The tissues were embedded and cut into 35-µm-thick cryosections. For immunofluorescence staining, sagittal cortex slices or cells were permeabilized (0.5% Triton X-100 and 3% BSA in PBS) for 15 min and blocked (0.3% Triton X-100 and 3% BSA in PBS) for 1 h at room temperature. Next, the slices or cells were incubated with relevant primary antibodies (0.3% Triton X-100 and 3% BSA in PBS) overnight at 4°C. The samples were incubated with AlexaFluor 488-, 568-, 594- or 647-conjugated secondary antibodies (1:500, Life Technologies), and nuclei were counterstained with DAPI (1:1000, Sigma). Antibody categories are listed in **Supplementary Table 2**.

### Microscopy and analysis of NPCs

Confocal images were acquired using Zeiss LSM880 confocal microscopes and analyzed by ZEN software.

### Mouse lines

The animal experiments were approved by the Laboratory Animal Center, Institute of Zoology, Chinese Academy of Sciences, and were performed in accordance with relevant Chinese regulations. *Mark2*^+/-^ mice were generated by Cyagen Biosciences (China) using double gRNA CRISPR/Cas-mediated genome engineering to delete exons 2-17 of *Mark2*. gRNA primers are listed in **Supplementary Table 1**. All mice were bred in a specific-pathogen-free facility on a 12 h light/dark cycle, and food and water were available ad libitum.

### BrdU incorporation analysis

BrdU (Sigma, B5002-5G) was administered by intraperitoneal injection at a dose of 100mg/kg body weight of the mother mouse. Brains were harvested 1 h later for subsequent analysis. Antibody categories are listed in **Supplementary Table 2**.

### Behavioral and memory tests

All mice used for the behavioral tests were male mice aged 8-12 weeks (*n*≥8 per group), and all tests were performed between 09:00 and 17:00. Videos of the behavioral tests were analyzed by EthoVision XT 14 (Noldus). Behavioral tests included open field, elevated plus maze, three-chamber, novel object recognition, marble-burying, grooming, Y-maze, Barnes maze and Morris water maze (see **Supplementary materials**).

### Statistical analysis

Data are presented as the mean±s.e.m. unless otherwise indicated. At least three biological replicates were performed for each group. For statistical analyses, unpaired Student’s t tests were performed using GraphPad Prism software. Statistical significance was defined as \**p*<0.05, \*\**p*<0.01, and \*\*\**p*<0.001.

### Data availability

The whole exome sequencing data will not be made publicly available, as they contain private information. The RNA-sequencing raw data and other analyses supporting the findings of this study are available from the corresponding authors upon request via https://ngdc.cncb.ac.cn/gsa-human.

## Results

### Variant spectrum and clinical features of *MARK2*-related ASD

We utilized the GeneMatcher data sharing platform^18^ to assemble this international cohort. A total of 29 distinct *MARK2* variants (**Fig. 1a and Supplementary Table 3**) were detected in 31 individuals, including two individuals previously reported in the Deciphering Developmental Disorders (DDD) study.^17^ The variants comprised 25 Lof variants (nonsense/frameshift/splicing) (81%) and six missense variants (19%). All Lof variants led to the loss of the entire KA1. Among these, 22 (71%) variants were *de novo*, and four (13%) were paternally inherited (**Table 1**). Notably, 29 (94%) variants were novel. Three individuals presented variants at the same position (Arg302), including two Lof variants (R302*) and one missense variant (R302Q). Two variants recurred in independent individuals from different ethnicities and laboratories (R302* in P8 and P12, Q747* in P20 and P24). No other clinically relevant or pathogenic variants were detected in all affected individuals. All variants were classified as likely pathogenic or pathogenic.^19^

**Figure 1.**
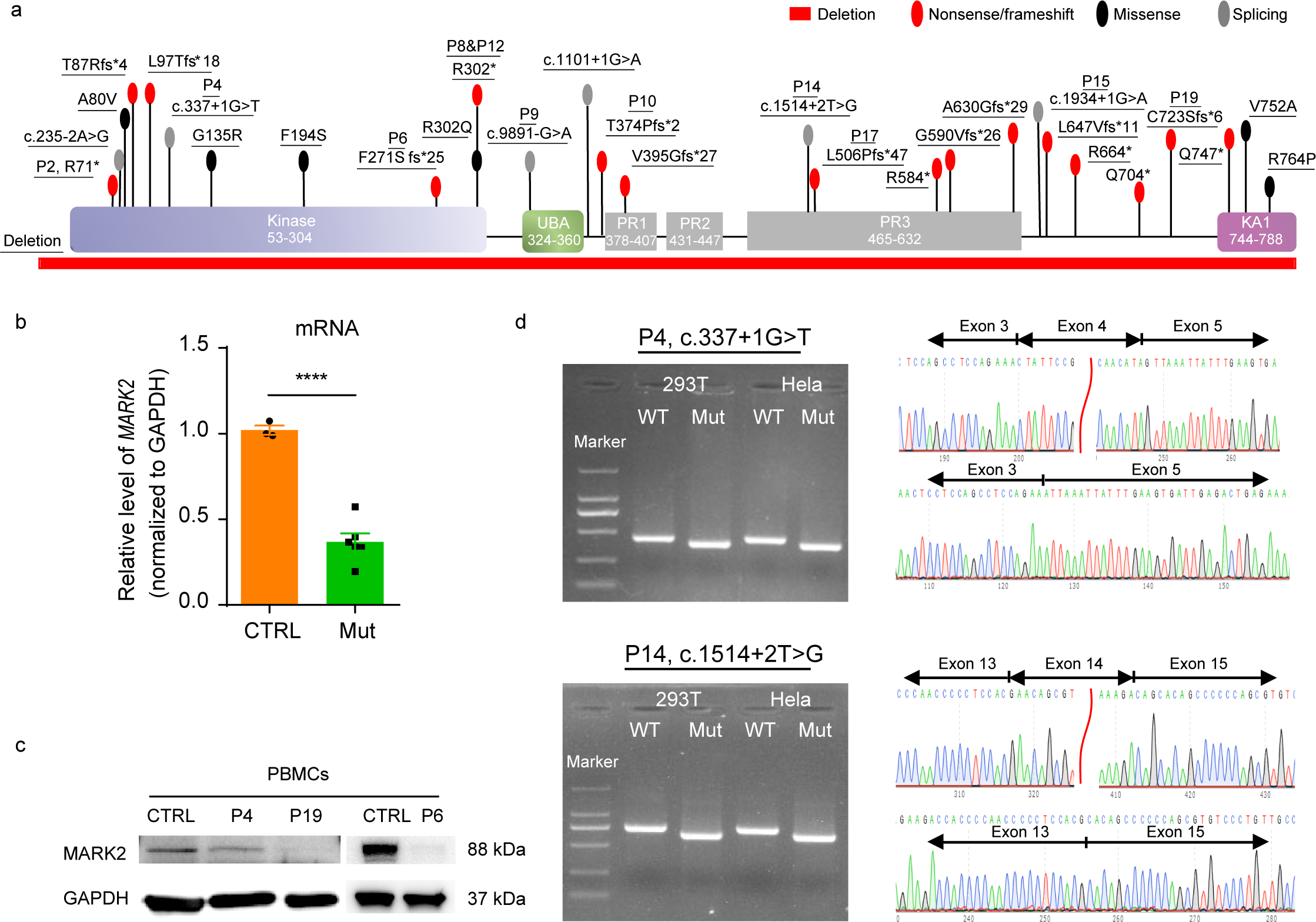
Clinical and genetic characteristics of referred individuals carrying *MARK2* variants. **a** Distribution of 29 variants of *MARK2* detected in 31 ASD individuals. R302* and Q747* are recurrent variants that occurred in two individuals. The locations of variants in functional domains of *MARK2* predicted by UniProt are shown. Kinase: protein kinase domain; UBA: ubiquitin-associated; PR: polar residue; KA1: kinase-associated domain 1. **b-c** The mRNA (**c**) and protein expression (**c**) of *MARK2* in the PBMCs from three affected individuals (P4, P6, P19). Both mRNA and protein levels were normalized to GAPDH levels and compared with those of age-matched healthy children (CTRL). The data are presented as the mean±s.e.m. of at least three independent experiments and were analyzed by Student’s t test, \**p*<0.05. **d** Minigene assays of two splicing variants (P4: c.337+1G>T; P14: c.1514+2T>G). RT□PCR products from HEK-293T and HeLa cells transfected with either the wild-type (WT) or mutant (Mut) pcDNA3.1 vector were separated by electrophoresis (**left panel**). M: DNA marker; the sizes of the bands are 2000, 1000, 750, 500, 250, and 100bp. Sequencing of the RT-PCR products (**right panel**) show c.337+1G>T variant results in a 49bp deletion, while the c.1514+2T>G variant results in a 98bp deletion.

**Table 1.**
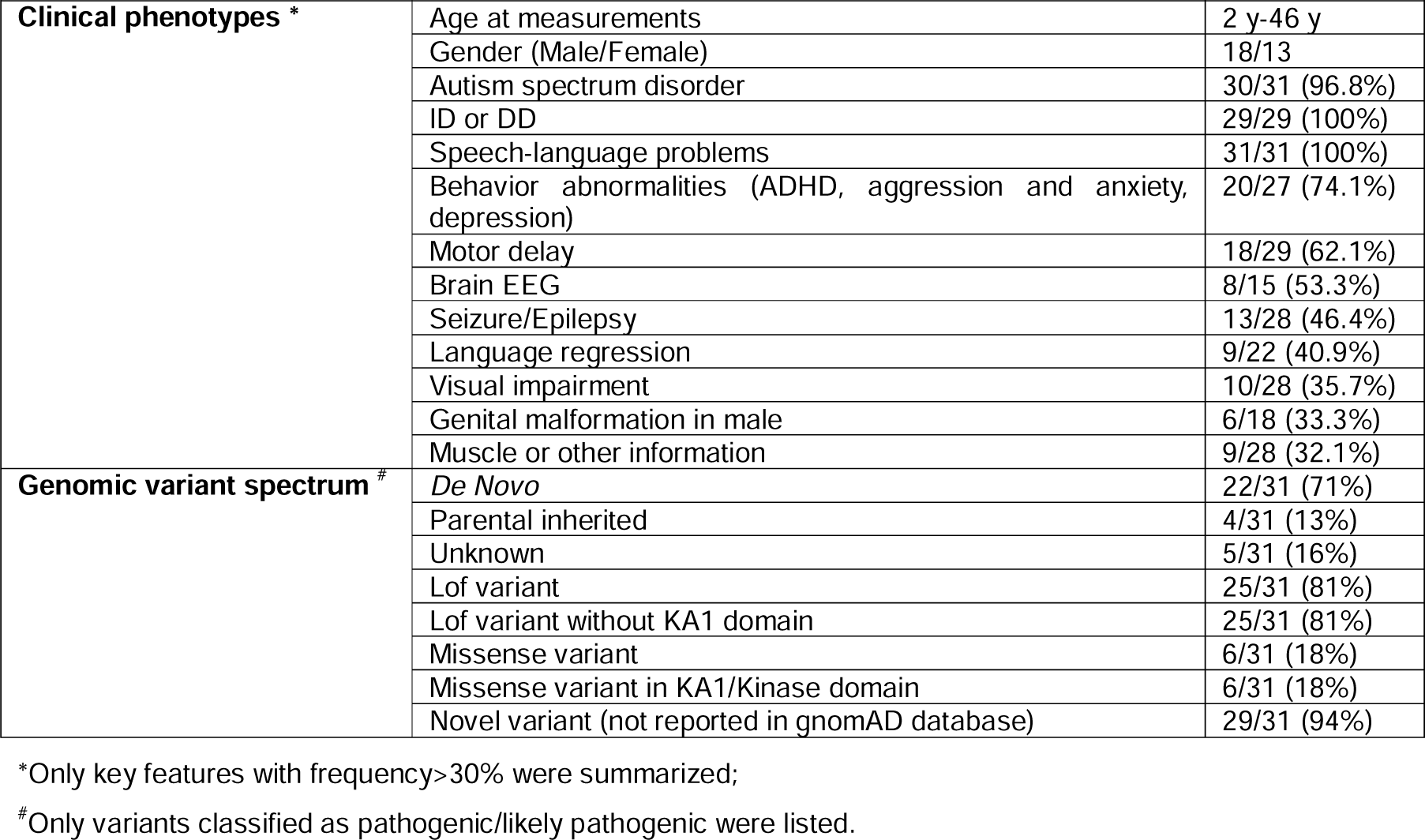
The detailed clinical features and variant spectrum of 31 independent individuals.

The detailed clinical features associated with the *MARK2* variants are summarized (**Supplementary Table 3**). The ages of individuals in our cohort ranged from 2 to 46 years old, with no observed sex difference (13 females and 18 males). Eight individuals displayed overlapping facial dysmorphisms, including round face, prominent forehead, hypertelorism, broad nasal root, large ears, thin upper lip and long philtrum. Eye defects, including hyperopia, myopia, astigmatism, strabismus, ptosis and visual processing difficulties, were observed in 46% individuals (11/24). All individuals except one (P8) presented with ASD (30/31). Other NDDs were common within the cohort, including ID/DD (29/29), speech-language problems (31/31), seizure/epilepsy (13/28), motor delay (18/29) and behavior disorders (20/27). Language regression was reported in 41% (9/22) of individuals with four individuals still presenting language regression (P6, P19, P8 and P9) over time. The high penetrance of NDDs suggested that *MARK2* is one candidate gene for syndromic and low-function ASD rather than isolated high-function ASD.

To assess the consequence of Lof variants, *MARK2* levels from PBMCs of three affected individuals (c.337+1G>T variant in P4, F271Sfs in P6, and C723Sfs variant in P19) were measured for both mRNA and protein *in vivo* (**Fig. 1bc**). As shown, the mRNA and protein expression of *MARK2* were significantly lower in affected individuals than in the control group (CTRL). We also validated the splicing effects of two canonical splicing variants (c.337+1G>T variant in P4 and c.1514+2T>G variant in P14) *in vitro* using a minigene assay. The transcripts of these *MARK2* variants (Mut) were shorter than that of wild-type (WT) one (**Fig. 1d**). Sequencing and online alignment of the RT-PCR products revealed one exon skipping event in the transcript, with a 49bp deletion in the c.337+1G>T variant and a 98bp deletion in the c.1514+2T>G variant. Both *in vivo* and *in intro* experiments confirmed *MARK2* loss due to Lof variants.

### Structural modeling and *in vitro* expression assay reveal destabilization caused by *MARK2* missense variants

MARK2 contains five domains, including kinase, KA1, ubiquitin-associated (UBA) and three polar residue domains.^9^ All missense variants identified in our cohort (A80V, G135R, F194S, R302Q, V752A, R764P) were clustered in the kinase or KA1 (**Fig. 1a**) domains and involved the highly evolutionary conserved residues across species from *Drosophila* to human (**Fig. 2a**). We analyzed the missense mutation tolerance map (https://stuart.radboudumc.nl/metadome/dashboard) by extracting missense and synonymous variants from the gnomAD database (gnomAD v.2.1.1, https://gnomad.broadinstitute.org/),^31^ and found both the kinase and KA1 domains are intolerant to missense mutations (**Fig. S1**).

**Figure 2.**
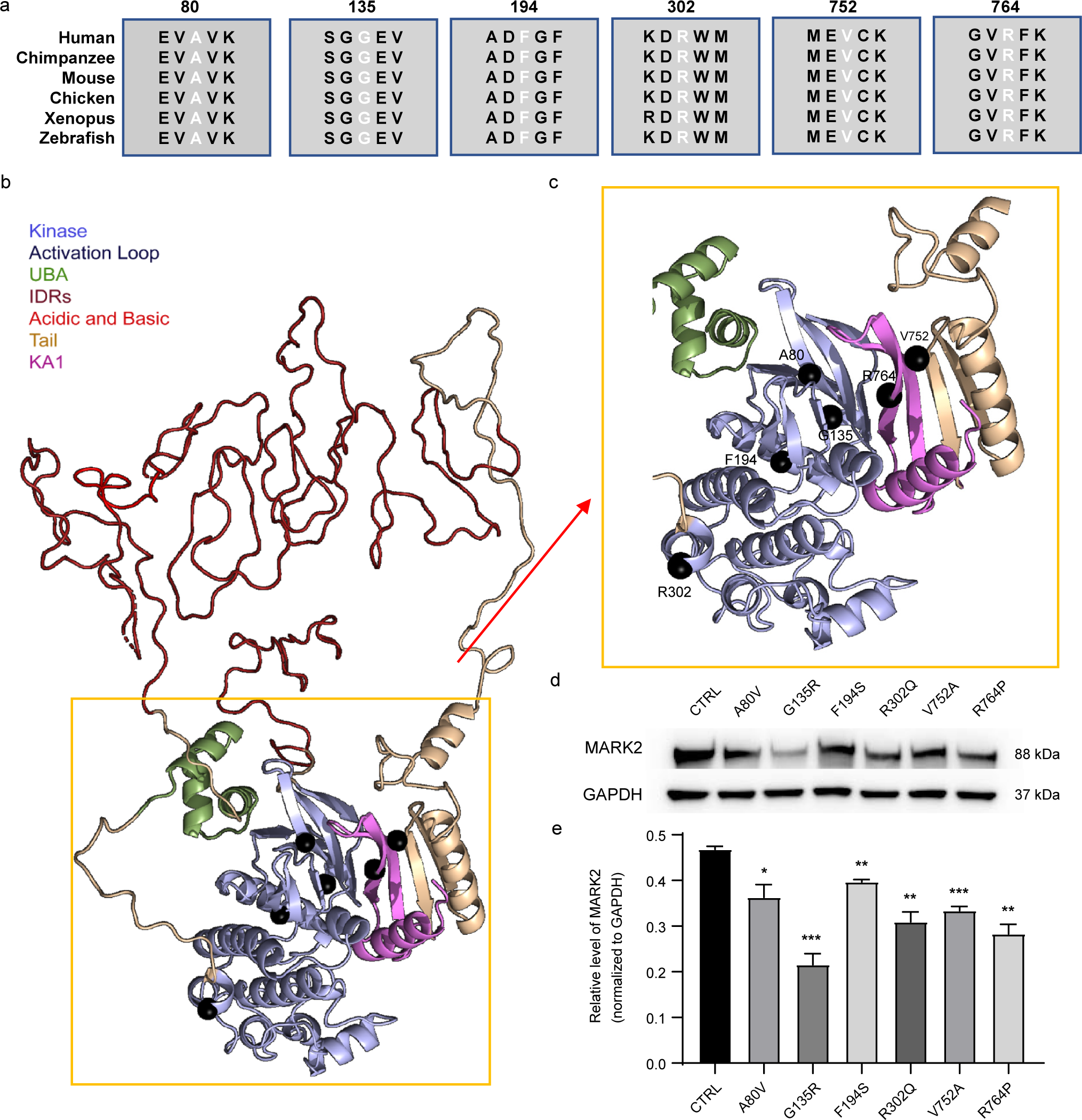
Structural modeling of six *MARK2* missense variants. **a** Alignment of the protein sequences for six *MARK2* missense variants across species from *Drosophila* to humans. **b** 3D structural models of *MARK2* missense variants. The protein domains of MARK2 are shown in different colors. The 3D structures of the non-intrinsically disordered regions (non-IDRs) were used to compute the changes in structural stability due to genomic variation. Amino acids in the IDRs are shown in a disordered configuration; we expect that any individual configuration of the IDRs would be an inadequate representation of their diverse dynamics. **c** The locations of the six missense variants in (**b**) are marked by black spheres. Four variants in the kinase domain (A80V, G135R, F194S, R302Q) and two variants in the KA1 domain (V752A, R764P) were all predicted to locate in the activation loop. **d-e** Representative western blot image (**d**) and quantification analysis of exogenous *MARK2* expression which were normalized by total GAPDH levels (**e**). Human HEK293T cells transfected with wild-type (CTRL) or mutant EGFP-*MARK2*-Neo vectors were lysed. The data are presented as the mean±s.e.m. of at least three independent experiments and were analyzed by Student’s t test \**p*<0.05, \*\**p*<0.01 and \*\*\**p*<0.001.

Information from evolutionary couplings indicates that the tail and KA1 domains fold together with the kinase domain (**Fig. 2b**),^32^ and six missense variants are within this structured dual domain (**Fig. 2c**). Amino acids in the kinase domain are expected to have a balanced frustration so that the kinase can move as part of the normal cycle of ATP/ADP exchange and substrate access. Further, the effects of six variants on stability, frustration, binding motifs, and biochemical role in the enzyme, the altered 3D domain structure or SLiMs were showed in **Supplementary Table 4** and **Supplementary Table 5**. The fraction of highly frustrated interactions around Ala80 is comparable, and the ΔΔG_fold_ of the A80V variant shows little to no change. However, the backbone of the Ala80 and Met129, the gatekeeper residues of the kinase fold, form a hydrogen bond, as they are in adjacent β-strands. The degree of freedom of Met129 may be restricted by the comparatively bulkier side chain of Val80, indirectly and negatively affecting the catalytic function via a dynamic mechanism. Both Gly134 and Gly135 are in the kinase domain, near the KA1 binding surface and solvent exposed. The ΔΔG_fold_ of Gly135Arg is highly unfavorable, suggesting that the variant destabilizes the kinase domain. Gly135 is also positioned in the ATP-binding pocket, and the introduction of a large polar amino acid to the vicinity is likely to dysregulate nucleoside binding. G135R variant introduces a new sequence motif for PCSK cleavage. If this new motif is recognized, it could represent a new manner in which MARK2 is inactivated. Phe194 is located in the N-terminal base of the kinase activation loop and is a key residue in the classic Asp-Phe-Gly (DFG) motif. Phe194 anchors the activation loop within the kinase domain by hydrophobic interaction, rendering optimum flexibility to the activation loop. Two new sequence motifs were gained in the F194S variant, one for phosphorylation by CK1 and another for glycosaminoglycan attachment. Thus, there is also potential for this genomic variant to introduce new regulatory dynamics into the enzyme, particularly given its proximity to known phosphorylation sites such as Ser197, Thr201, and Thr208. Arg302 is in the C-terminal helix of the kinase domain, which connects to the UBA domain. Arg302 was solvent exposed in our model and formed a hydrogen bond network among the Asp276 residue of the kinase, the N-terminal loop connected to the UBA domain, and additional N-terminal residues of the kinase. The frustration level indicated that R302Q variant resulted in destabilization, while the ΔΔG_fold_ indicated that this variant caused stabilization. According to structural analysis, the hydrogen bond between Gln302 and Asp276 was preserved in the mutated protein. Thus, R302Q variant can be tolerated but may also have allosteric effects on the kinase, as we have previously reported.^33^ Val752 residue is in the core of the hydrophobic interaction between the KA1 and tail domains. V752A is a cavity-forming variant that may destabilize the hydrophobic core, potentially disrupting the formation of this autoregulatory domain. Similar to the kinase domain mutations that destabilize KA1 interactions, R764P is part of the same interaction region and therefore is a reciprocal change with similar instabilities induced in the kinase-autoregulatory interaction. The ΔΔG_fold_ value suggested that V752A variant is destabilizing, leaving a weaker autoinhibition state. The six missense variants all result in changes that may result in loss of kinase catalytic function, we then hypothesize that the missense alterations observed in this cohort could lead to *MARK2* destabilization or inactivation.

In order to test this hypothesis, we constructed and transfected pCMV-Neo-MARK2 vectors into 293HEK cells, and analyzed the exogenous *MARK2* expression. As shown in **Fig. 2de**, six mutant cells resulted in significantly lower *MARK2* expression compared with WT cells. Among them, G135R had the lowest *MARK2* expression. These *in vitro* data suggested that missense variants in the kinase and KA1 domain can result in decreased protein expression due to *MARK2* destabilization, and thereby phenocopy *MARK2* loss arising from Lof variants.

### *MARK2* variant leads to anomalous polarity in iPSC-derived neural rosettes and the imbalanced proliferation and differentiation in iPSC-derived NPCs

Although the roles of *MARK2* plays in neural development are recognized, such as hippocampal neuronal polarity and migration, dendritic development,^11–16^ the cellular development phenotypes and fundamental mechanism of *MARK2* variant or loss during early neural development are unclear. To clarify it, we generated iPSCs from the PBMCs of two affected individuals (P19:C723Sfs; P6:F271Sfs), two healthy males (CTRL1 and CTRL2) (**Fig. 1a and S2a**), and two isogenic mutant iPSCs (CRISPR-Del1 and CRISPR-Del2). These six iPSCs, with two or more clones for each iPSCs, were generated successfully. Their pluripotencies (SOX2, OCT3/4, and NANOG, **Fig. S2b**), karyotypes (**Fig. S2a**) and differentiation abilities to the three germ layers were assessed.

Neural rosettes develop from the self-organized and differentiated iPSCs and present radial arrangements of neuroepithelial cells with one central lumen resembling developing neural tube.^34^ The key morphology of neural rosette includes intercalation, constriction, polarization, elongation and lumen formation.^34, 35^ We compared the morphology of iPSC-derived neural rosettes at different developmental stages. Firstly, we found that both mutant and CRISPR-Del neural rosettes were different from CTRLs (**Fig. 3abd, yellow dotted loop**), with the number of rosettes per 100 cells being significantly decreased (**Fig. 3g**) and the diameters of the rosettes being significantly increased (**Fig. 3f**), suggesting few and unstable, loose neural rosette formation after *MARK2* loss. Considering the role of *MARK2* in neural polarity,^11^ we furtherly co-stained neural rosettes using specific antibodies for neuronal marker (TUJ1) and polarity marker (ZO1) (**Fig. 3a**). CTRL rosettes exhibited well-defined self-organization characterized by apical-basal polarity and constriction. TUJ1 staining revealed radial distribution surrounding the inner lumen, while ZO1 staining demonstrated clear localization within the inner lumen, which is consistent with previous reports.^35^ In contrast, both mutant (C723Sfs and F271Sfs) and CRISPR-Del rosettes exhibited loosely structure, characterized by imperfectly self-organized TUJ1 staining and degraded ZO1 staining, along with the absence of an inner lumen (**Fig. 3a**). These results demonstrated that *MARK2* loss led to aberrant polarity of neuroepithelial cells, forming dis-organized, few and loose-structured neural rosettes. Further comparation of the diameters of different iPSC-derived neurospheres revealed a significant reduction in size in mutant and CRISPR-Del groups (**Fig. 3ci**). We then checked the undifferentiated cell pool in rosettes using SOX1.^36^ Also, we observed a significant decrease in the number of positive SOX1^+^ cells within mutant and isogenic neural rosettes, indicating aberrant self-renewal of rosette following *MARK2* loss (**Fig. 3bh**).

**Figure 3.**
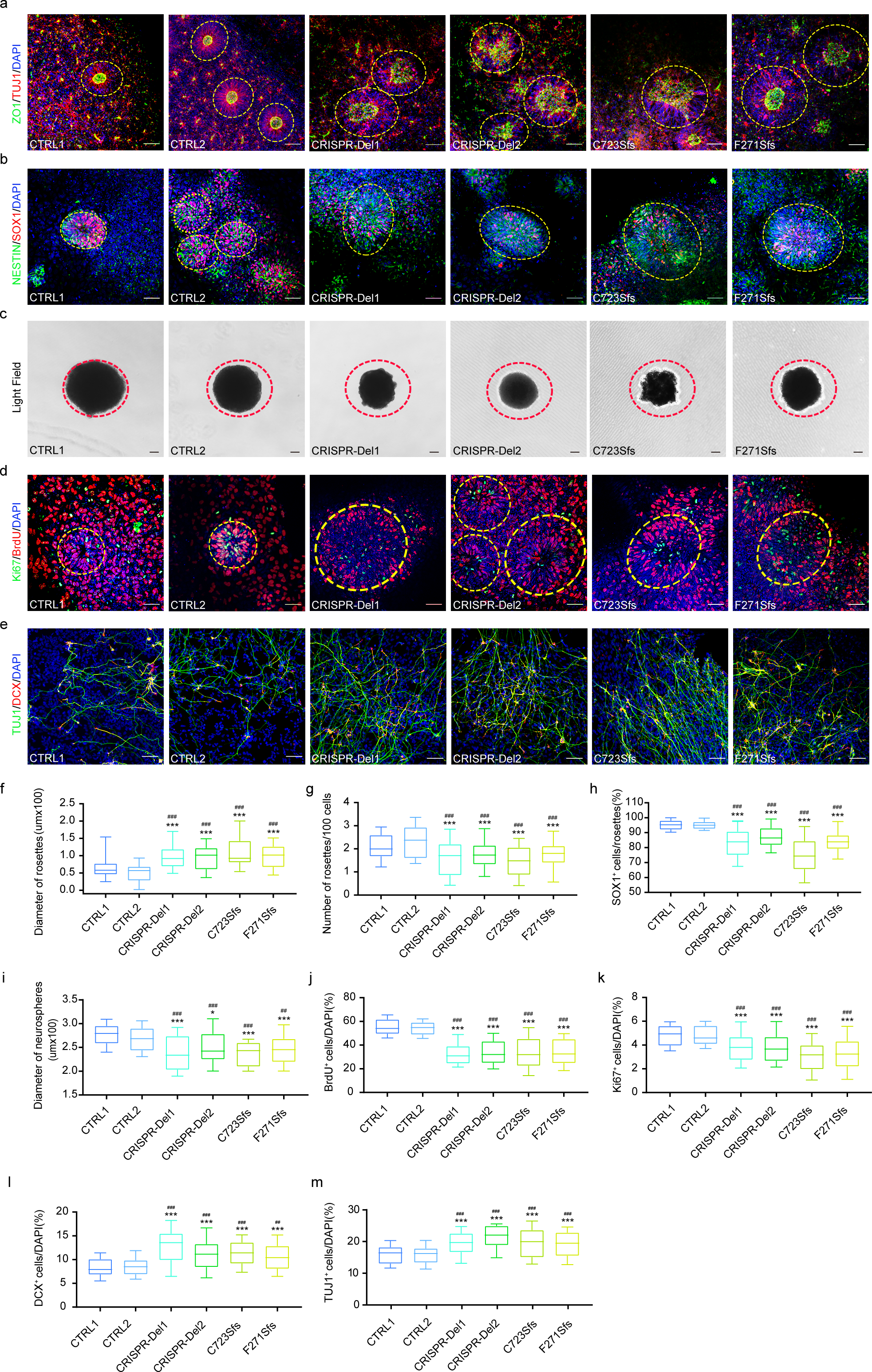
*MARK2* variant leads to aberrant polarity in iPSC-derived neural rosettes and imbalanced proliferation and differentiation in iPSC-derived NPCs. **a** Representative immunofluorescence images of TUJ1 (red) and ZO1 (green) in different iPSC-derived neural rosettes on the 10th day after neural induction. CTRL1 and CTRL2: two independent healthy adults without *MARK2* variant; C723Sfs and F271Sfs: two affected individuals with *MARK2* variant; CRISPR-Del1 and CRISPR-Del2: two isogenic *MARK2* deletion produced by the CRISPR/Cas9 editing technology. **b** Representative immunofluorescence images of SOX1 (red) and NESTIN (green) in different iPSC-derived neural rosettes on the 5th day after neural induction. **c** Representative images of different iPSC-derived neurospheres on the 5th day after neural induction. **d** Representative immunofluorescence images of BrdU (red) and Ki67 (green) in different iPSC-derived neural rosettes on the 4th day after neural induction. **e** Representative immunofluorescence images of DCX (red) and TUJ1 (green) in different iPSC-derived NPCs on the 13th day after neural induction. **f-m** the quantification analysis of rosette diameter (**f**, *n*=12 rosettes), rosette number (**g**) in panel a, SOX1^+^ cells (**h**) in panel b, neurosphere diameter (**i**, *n*=12 neurospheres) in panel c, BrdU^+^ cells (**j**) and Ki67^+^ cells (**k**) in panel D, DCX^+^ cells (**l**) and TUJ1^+^ cells (**m**) in panel E. The data are presented as the mean±s.e.m. of at least three independent experiments and were analyzed by Student’s t test; \**p*<0.05 and \*\*\**p*<0.001(compared with CTRL1); ^##^*p*<0.01 and ^###^*p*<0.001(compared with CTRL2). scale bar = 50 μm.

In iPSC-derived NPC stage, we conducted BrdU incorporation assays and Ki67 immunofluorescence (IF) staining on to assess proliferation capability (on the 4th day after differentiation induction, **Fig. 3d**). The results demonstrated that there were significantly less BrdU^+^ cells (**Fig. 3j**) and Ki67^+^ cells (**Fig. 3k**) in mutant or CRISPR-Del NPCs compared to CTRLs While the proliferation and differentiation of NPCs maintain a dynamic balance, we also evaluated the differentiation efficiency of NPCs on the 13th day after differentiation induction using two neural markers, TUJ1 and DCX (**Fig. 3e**). TUJ1^+^ cell (**Fig. 3l**) and DCX^+^ cells (**Fig. 3m**) in mutant and CRISPR-Del NPCs also showed a significant increase compared to CTRLs. Together, these results suggested that *MARK2* loss led to aberrant polarity of neuroepithelial cells, dis-organization of neural rosette, thereby suppressing proliferation but promoting differentiation in iPSC-derived NPCs.

### *Mark2* loss in mice lead to abnormal cortical development, ASD-like behaviors and impaired memory

To recapitulate the cellular phenotypes of patient’s iPSC-derived rosettes, we generated *Mark2*^+/-^ (HET) mice utilizing the CRISPR/Cas9 system (**Fig. S3a S3d**) to mimic *Mark2* loss *in vivo*. Consistently, the BrdU incorporation assay of mouse cortical NPCs (mNPCs) at E18.5 revealed significantly decreased BrdU^+^ cells but increased TUJ1^+^ cells in HET mice compared with wild-type (WT) ones (**Fig. S3eh**), indicating the imbalanced proliferation and differentiation in early cortical NPCs upon *Mark2* loss. Additionally, downregulated SOX1^+^ cells were also observed in the cortices of HET mice (**Fig. S3eg**) indicating advanced/altered neural differentiation upon *Mark2* loss. Using Ctip2 as a marker for the middle cortex region (layers II/III and V) and Stab2 as a marker for the whole cortex,^37^ we compared the thickness of the different cortex layers between WT and HET mice (**Fig. S3f**). The Ctip2^+^ and Stab2^+^ layers in HET mice were significantly thicker than those in WT mice (**Fig. S3jk**). These results indicated abnormal cortical development and partitioning upon *Mark2* loss.

In order to determine whether adult *Mark2*^+/-^ mice exhibit the features of ASD and other NDDs observed in affected individuals, we performed series of behavioral tests for *Mark2*^+/-^ mice. Although HET mice had lower body weights (**Fig. S3c**), they showed normal exploratory and locomotor activity in the open field test (**Fig. S4a**), as the total distance traveled (**Fig. S4c**) and average speed (**Fig. S4d**) were not different between HET mice and WT mice. Subsequently, we performed the three-chamber test to evaluate whether the mice exhibited the deficits in sociability and preference for novelty observed in ASD patients.^38^ In the social approach test (**Fig. 4ab**), HET mice spent similar amounts of time in the empty cage as WT mice but spent significantly less time in the cage containing the novel mice than WT mice. In the social novelty test (**Fig. 4ef**), HET mice spent similar amounts of time as WT mice with the familiar mice but spent significantly less time with the novel mice. These social behavior test demonstrated that HET mice presented reduced social motivation and pursuit of novelty. The marble burying test and grooming test were used to evaluate stereotyped behaviors of ASD in rodents. We found that HET mice buried more marbles (**Fig. 4cd**) and spent more time grooming (**Fig. 4k**) than WT mice. Additionally, in the elevated plus maze test, which was used to assess anxiety-like behavior of ASD, HET mice spent less time in the open arms than WT mice (**Fig. 4hi**). Together, these data suggested that *Mark2* loss in mice led to specific social deficits, stereotyped behavior, and anxiety that recapitulated the features of ASD in individuals carrying *MARK2* variants.

**Figure 4.**
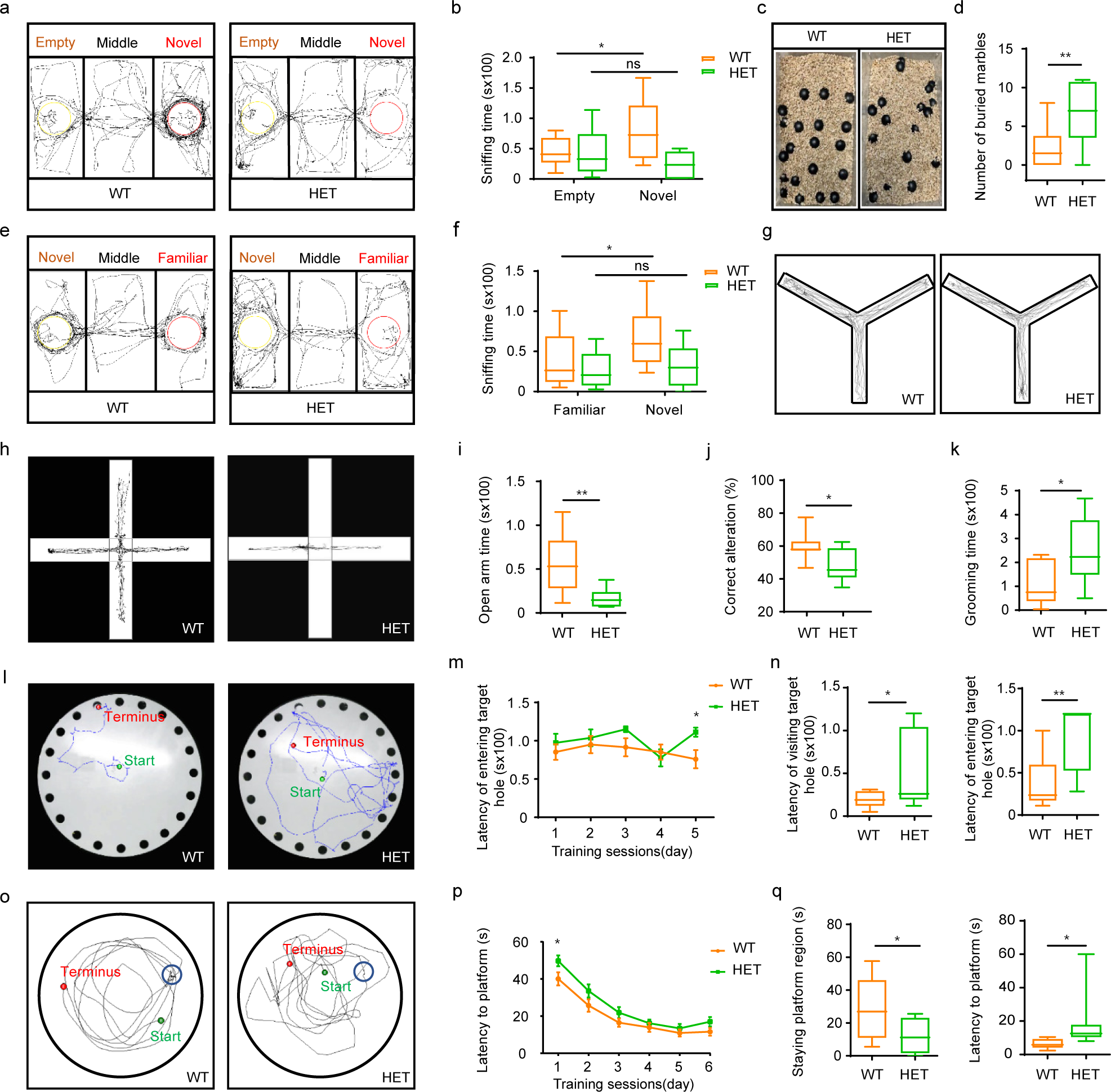
*Mark2* loss in mice leads to ASD-like behavior and impaired memory. **a&e** Trajectories of *Mark2* ^+/+^ (WT) and *Mark2* ^+/-^ (HET) mice in the three-chamber test. **b&f** Quantification analysis of sniffing time in the three-chamber test (**b**, empty or novel; **f**, novel or familiar). **c-d, k** Trajectories of WT and HET mice in the marble-burying test (**c**) and quantification analysis of the buried marbles (**d**) and grooming time (**k**). **h-i** Trajectories of WT and HET mice in the elevated plus maze test (**h**) and quantification analysis of time spent in the open arms (**i**). **g&j** Trajectories of WT and HET mice in the Y-maze test (**g**) and quantification analysis of the correct alternation (**j**). **l-n** Trajectories of WT and HET mice in the Barnes maze test (**l**), quantification analysis of the latency of entering the target hole during the training sessions (**m**), the latency of visiting target hole and of entering target hole during the test session (**n**). **o-q** Trajectories of WT and HET mice in the Morris water maze test (**o**), quantification analysis of the latency to find the platform during the training sessions (**p**), the latency to finding the platform and staying platform quadrant during the test session (**q**). WT=9, HET=9. The data are expressed as the mean±s.e.m. and were analyzed by Student’s t test; \**p*<0.05 and \*\**p*<0.01.

As affected individuals in our cohort also showed ID/DD and language problem, we further assessed learning and memory capacity using the Y-maze (**Fig. 4g**), the Barnes maze (**Fig. 4l**), and the Morris water maze (**Fig. 4o**). The Y-maze is designed to assess spatial memory and executive function. We found that the percentage of correct alterations made by HET mice was significantly decreased compared with WT mice (**Fig. 4j**), indicating that HET mice exhibited abnormal spatial working memory. The results of the Barnes maze test suggested memory impair in HET mice, as they spent significantly more time visiting (**Fig. 4n left**) and entering (**Fig. 4m, 4n right**) the target hole compared with WT ones. In the Morris water maze test (**Fig. 4o**), HET mice showed a significantly longer latency to find the platform (**Fig. 4p,4q right**) and spent less time in the platform region when the platform was moved (**Fig. 4q left**). Interestingly, during the training sessions of the Barnes maze and Morris water maze tests, the latencies of the HET mice were not significantly longer than those of the WT mice, suggesting similar learning abilities between the two genotypes. Nonetheless, we tested the recognition memory of mice using the novel object recognition test (**Fig. S4b**) and found that HET mice spent similar amounts of time interacting with the novel object as WT mice (**Fig. S4e**). Combined, these results implied that loss of *Mark2* impaired spatial memory.

### *MARK2* loss disrupts early neural development via the downregulation of WNT/**β**-catenin signaling pathway

To further identify the molecular pathway of the *MARK2* variant in early neurogenesis, we performed RNA-Seq for iPSC-derived NPCs on the 12th day after differentiation induction (**Fig.S5a**) to detect significant downregulated genes and upregulated genes (**Fig.S5b**, p. adjust>0.05, Log2Fold change>1.5). Gene Ontology (GO) enrichment analysis of DEGs revealed the biological functional change related to muscle development, ear development, neuro fate commitment/specification, axonogenesis, synapse development, or material transport with *MARK2* loss (**Fig. S5c**). GO enrichment analysis of downregulated genes are significantly involved in neural development (**Fig. 5a**), including central nervous system neuron differentiation (GO:0021953), forebrain development (GO:0030900), axonogenesis (GO:0007409), axon development (GO:0061564), pattern specification process (GO:0007389), negative regulation of neuron differentiation (GO:0045665), regulation of neuron differentiation (GO:0045664), regionalization (GO:0003002), axon guidance (GO:0007411), and neuron projection guidance (GO:0097485). Combined gene network analysis further demonstrated that WNT3A involved in all these GO terms, especially the terms related to early neural development (**Fig. 5b**). GSEA analysis also revealed significant inhibition of the WNT signaling pathway in mutant cells (**Fig. 5c**). Meanwhile, both mutant and CRISPR-Del cells exhibited significantly decreased WNT3A (**Fig. 5df**) along with decreased β-catenin (**Fig. 5de**). A previous study showed that WNT3A, as a critical ligand, plays an important role in the activation of the WNT signaling pathway, and is essential for the proliferation and differentiation of NPCs.^39^ Hence, we hypothesized that *MARK2* loss led to abnormal early neural development via inhibiting the WNT signaling pathway.

**Figure 5.**
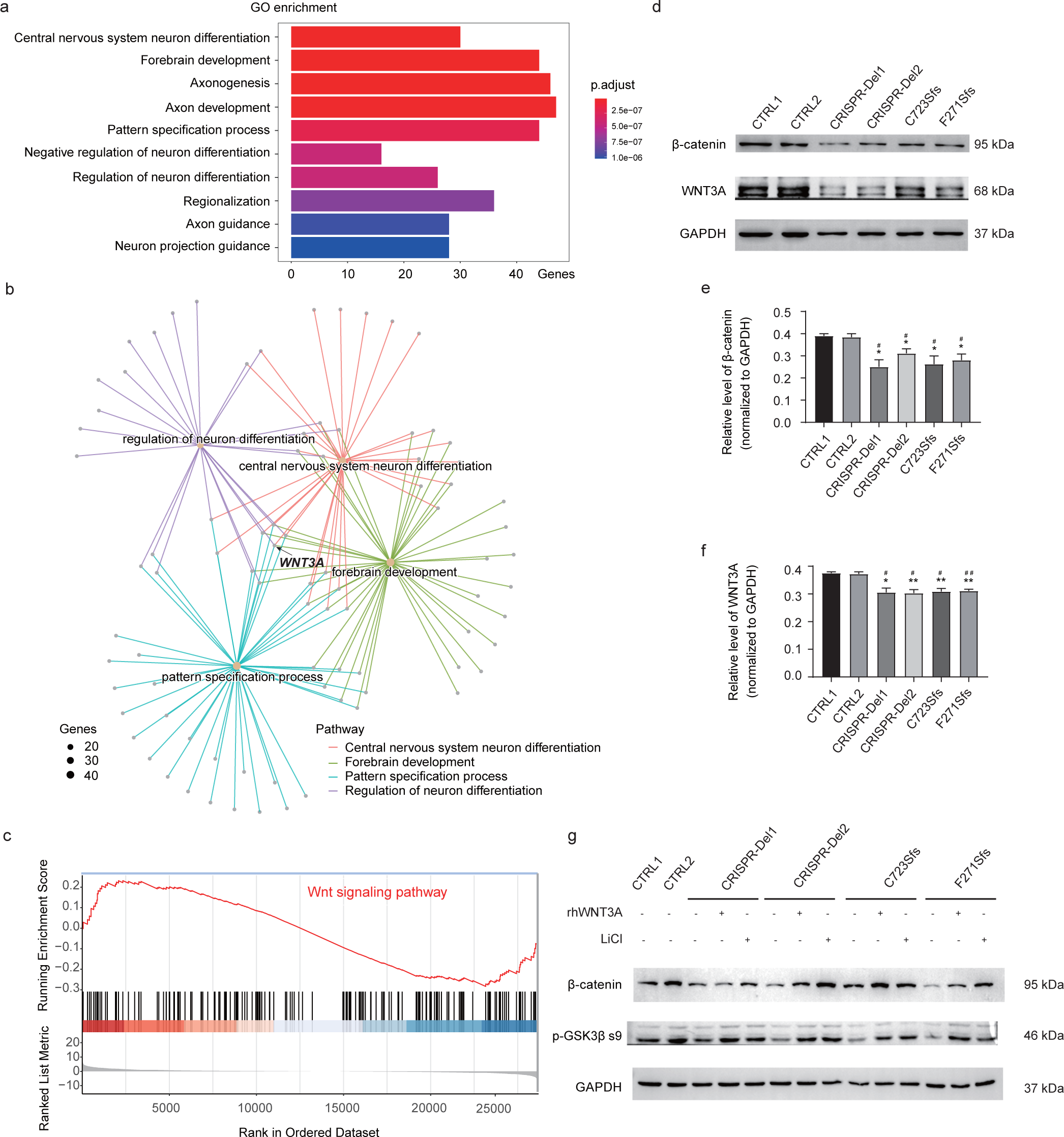
iPSC-derived NPCs revealed decreased WNT signaling pathway due to *MARK2* loss. **a** Gene ontology (GO) analysis of differentially expressed genes (DEGs) between mutant iPSC-derived NPCs and CTRL ones show that the downregulated genes are enriched in multiple pathways related to early neuronal development. **b** GO pathway network analysis showed the association with neuronal development, including neuron fate specification, forebrain development, central nervous system neuron differentiation, axonogenesis and axon development. Size means number of genes involved in the specific pathway. **c** The enrichment score and rank of the WNT/β-catenin signaling pathway from DEGs. **d-f** Representative western blot images and quantification analysis of β-catenin and WNT3A expression in mutant and CRISPR-Del iPSC-derived NPCs compared with CTRLs. **g** Representative western blot images and quantification analysis of β-catenin and p-GSK3β-s9 expression in mutant, CRISPR-Del iPSC-derived NPCs with/without rhWNT3A or LiCI treatment. The data are presented as the mean±s.e.m. of at least three independent experiments and were analyzed by Student’s t test; \**p*<0.05 and \*\**p*<0.01(compared with CTRL1); ^#^*p*<0.05 and ^##^*p*<0.01(compared with CTRL2).

In order to confirm the relationship between WNT signal pathway inhibition and *MARK2* loss, we treated mutant iPSC-NPCs with 100mg/ml rhWNT3A. We found that rhWNT3A significantly increased the expression of p-GSK3β s9, and the expression of β-catenin in mutant iPSC-NPCs (**Fig. 5g**). The rescue effect of rhWNT3A confirmed that *MARK2* loss results in relative inactivation of WNT signaling pathway.

### Lithium reverses the molecular and cellular phenotypes of mutant iPSC-derived NPCs and abnormal cortical partition development of HET mice by activating the WNT/**β**-catenin signaling pathway

Lithium is a known activator of the WNT/β-catenin signaling pathway^40, 41^ and is widely prescribed for many behavioral disorders, such as bipolar disorder and schizoaffective disorder,^42^ which are closely associated with ASD. We added LiCl at 0.7 mM, a routinely prescribed dosage for patients with bipolar disorder to the culture medium of iPSC-derived NPCs.^42, 43^ On the 10th day after differentiation induction, the protein levels of β-catenin, p-GSK3β s9 in mutant NPCs were significantly increased following LiCl treatment as similar to rhWNT3A treatment, even reaching the levels of control NPCs (**Fig. 5g**), suggesting WNT3A and p-GSK3β s9 as the target molecular of lithium rescue.

Considering the molecular effect of LiCl on WNT/β-catenin signaling activation, we replicated the IF experiment described above for LiCl-treated and untreated mutant and CRISPR-Del iPSC-derived NPCs. Innovatively, the rescue effect of LiCl on the morphology of mutant neural rosette was quite obvious, with the diameter of the neural rosettes being significantly decreased (**Fig. 6ad**) and the number of rosettes being significantly increased (**Fig. 6ae**). Also, correct localization of ZO1 and normal self-organization of TUJ1^+^ cells were seen in mutant neural rosettes with LiCl treatment. Moreover, the numbers of BrdU^+^ (**Fig. 6bf**) and Ki67^+^ cells (**Fig. 6bg**) in mutant and CRISPR-Del NPCs were significantly increased, almost reaching those of control NPCs. We further compared the differentiation efficiencies of mutant NPCs before and after LiCl treatment. The numbers of TUJ1^+^ (**Fig. 6ci**) and DCX^+^ cells (**Fig. 6ch**) were significantly decreased after LiCl treatment. Both molecular and cellular phenotypes revealed that lithium rescues the abnormal developmental trajectories of *MARK2* loss via stimulating the WNT signaling pathway.

**Figure 6.**
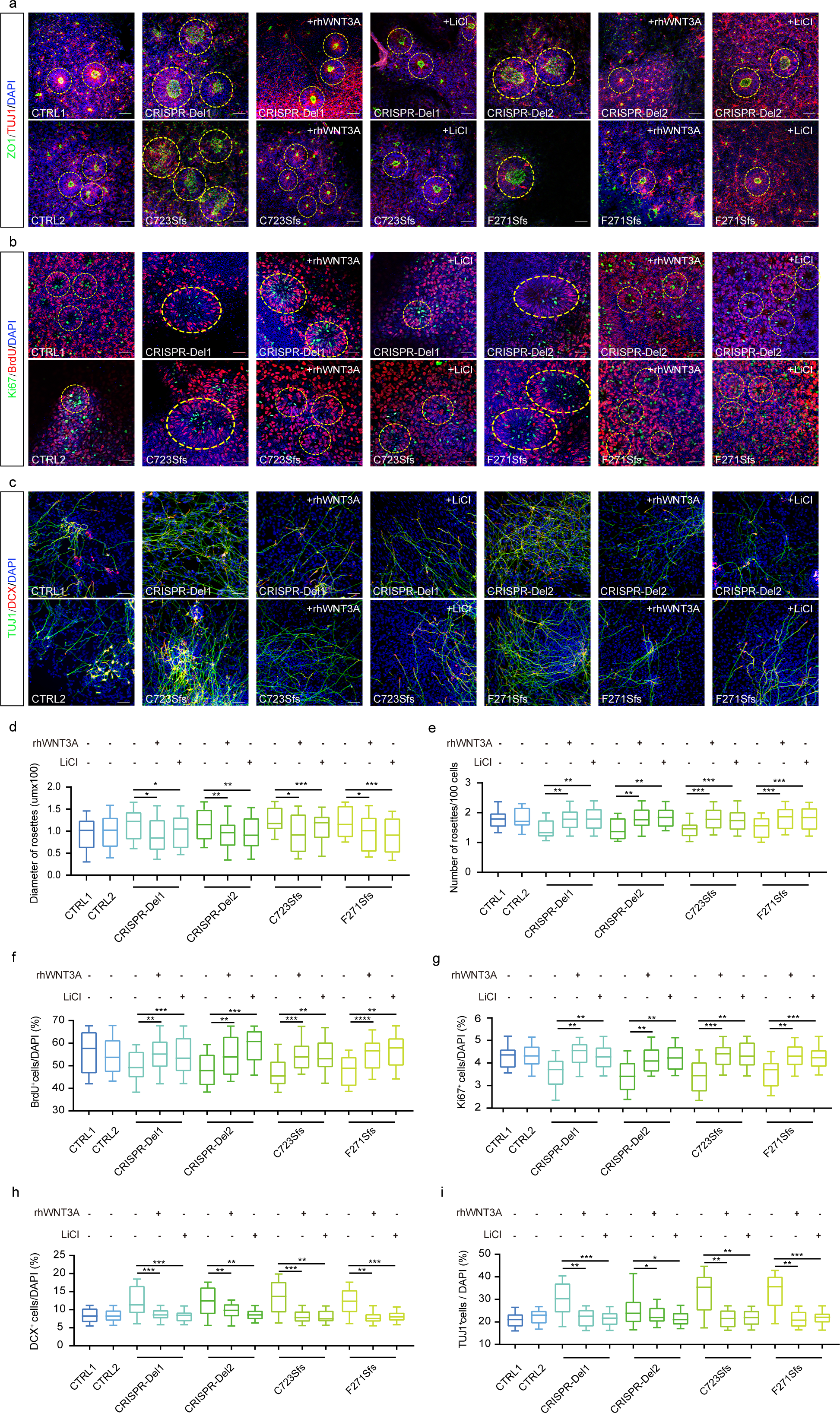
Lithium reverses the imbalance in the proliferation and differentiation of iPSC-derived NPCs with *MARK2* mutation by activating the WNT/β-catenin signaling pathway. **a** Representative immunofluorescence images of ZO1 (green) and TUJ1 (red) in different iPSC-derived neural rosettes. CTRL1 and CTRL2: two independent healthy adults without *MARK2* variant; C723Sfs and F271Sfs: two affected individuals with *MARK2* variant; CRISPR-Del1 and CRISPR-Del2: two isogenic *MARK2* deletion produced by the CRISPR/Cas9 editing technology. Both CRISPR-Del and mutant iPSC-derived neural rosettes were treated with LiCl (+LiCI) or rhWNT3A (+rhWNT3A). **b** Representative immunofluorescence images of Ki67 (green) and BrdU (red) in different iPSC-derived NPCs. **c** Representative immunofluorescence images of TUJ1 (green) and DCX (red) in different iPSC-derived NPCs. **d-i** The quantification analysis of rosette diameter (**d**), rosettes number (**e**) in panel A, BrdU^+^ cells (**f**) and Ki67^+^ cells (**g**) in panel B, DCX^+^ cells (**h**) and TUJ1^+^ cells (**i**) in panel C. The data are presented as the mean±s.e.m. of at least three independent experiments and were analyzed by Student’s t test; \**p*<0.05, \*\**p*<0.01 and \*\*\**p*<0.001. scale bar = 50 μm.

To investigate the rescue effect of LiCl on cortical layer development in *Mark2*^+/-^ mice, we treated pregnant female mice with 4.5 mg/L LiCl from E0.5 to E17.5 and analyzed proliferation and differentiation efficiency in the cortex at E18.5. We found that the number of BrdU^+^ cells (**Fig. S6ac**) was significantly increased in HET mice treated with LiCl, but the number of TUJ1^+^ cells was significantly decreased (**Fig. S6ae**). An obvious increase in the number of SOX1^+^ cells was also observed in LiCl-treated HET mice compared with untreated HET mice (**Fig. S6ad**). These data demonstrated the rescue effects of lithium on the proliferation and self-renewal of NPCs in mice with *Mark2* loss. We further compared the thickness of the cortical layers in HET mice before and after LiCl treatment using Ctip2 and Stab2 as markers of different cortex layers (**Fig. S6b**). The results showed that LiCl treatment significantly rescued the abnormal formation of the cortical layers, as the thickness of the Ctip2^+^ and Stab2^+^ cortical layers was similar between LiCl-treated HET mice and WT mice (**Fig. S6fg**). These results were consistent with the rescue effects observed in Mut iPSC-derived NPCs.

Together, these *in vitro* and *in vivo* results demonstrated that pharmacological reactivation of the WNT/β-catenin signaling pathway by lithium can rescue aberrant cellular phenotypes in NPC with *MARK2* loss, thereby indicating a potential molecular link between *MARK2* loss and WNT/β-catenin signaling.

## Discussion

In our study, we identified a multi-institutional global cohort of 31 individuals carrying clinically relevant *MARK2* variants, and defined a comprehensive phenotypic profile of *MARK2*-related ASD. Zhou et al analyzed the cognitive function of seven ASD individuals carrying Lof variants of *MARK2* and found that their average IQ (65.2) was similar to that of ASD individuals carrying *CHD8/SHANK3* variants, defining that *MARK2* is a high-confidence but low-function ASD gene.^17^ *MARK2* also has been identified as new candidate gene for NDDs based on *de novo* CNV enrichment.^44^ In our cohort, we also observed high frequency of comorbidities including ID/DD (100%), speech-language problems (29/29) and motor delay (61%). We also noted that one individual in our cohort (P8) has not meet ASD criteria till her last interview, although she presented mixed receptive-expressive language disorder (personal communication), implying *MARK2* is as the candidate gene of NDDs. Moreover, affected individuals exhibit typical facial dysmorphism (65%), including a prominent forehead, a broad nasal root, larger ears, a thin upper lip, and a long philtrum, visual problem, which may help us to identify additional *MARK2*-related ASD individuals. Eighty percent of *MARK2* variants are Lof variants, supporting the idea that *MARK2* haploinsufficiency contributes to the development of ASD, as initially suggested from two large sequencing studies.^17, 44^

MARK2 contains five domains, each with distinct functions necessary for its normal activity.^9^ Previous studies have reported that *MARK2* mutants lacking the kinase domain led to neuronal polarity abrogation in hippocampal neurons.^11^ Furthermore, mutations in the activating loop of the kinase domain, T208A and S212A, result in abnormal differentiation and neurite extension in neuroblastoma cells,^15^ suggesting the critical role of the kinase domain in neural development. The KA1 domain, locating in the C-terminus of *MARK2*, is conserved from yeast to humans and exerts an autoinhibitory effect on the kinase activity of *MARK2* by blocking peptide substrate binding to the N- and C-lobes of the kinase domain.^10^ Mutation in the KA1 domain, impairs this autoinhibition of MARK/PAR1 kinase activity.^32^ We identified four variants in the ATP-binding pocket or the activation loop of kinase domain, and two variants in the autoregulatory position of KA1 domain. Using integrative bioinformatics approaches, we predicted that these variants destabilize the interaction between these two domains and lead to loss of the autoregulation of *MARK2*. Further, our *in vitro* transfected cell model confirmed that missense variants result in decreased *MARK2* expression comparable to Lof variants. Additionally, the phenotype associated with the missense variants was not significantly different from that of Lof variants. These data suggest that the missense variants impair the kinase or autoinhibitory activity of MARK2 following protein destabilization resulting in similar cellular and molecular consequences as *MARK2* loss.

Multiple neurodevelopmental processes, including proliferation/neurogenesis, migration, neurite outgrowth, morphogenesis of dendrites and dendritic spines, and synaptogenesis and gliogenesis, have been reported to be involved in ASD.^45–48^. Although the pathophysiological mechanism by which *MARK2* loss underlies ASD in humans is unknown, the role of *MARK2* in neuronal development is recognized,^11–16^ including mediation of neurite outgrowth,^11, 12, 15^ establishment of neuronal polarity,^11, 12, 15^ specification of neuronal dendrites/axons,^11, 12, 16^ and promotion of neuronal migration.^13, 14^ In our study, we obtained mutant human iPSCs, generated *Mark2*^+/-^ mice, and explored the cellular abnormalities during the early neural developmental processes and its molecular pathways. For the first time, cellular phenotypes of mutant neurons at early neural development were demonstrated, such as less neural rosettes with larger diameters, depolarized and loose structure, smaller neurosphere formation, imbalanced proliferation and differentiation in NPC stage, which were similar to previous iPSC studies of ASD.^46–48^ Beside *MARK2*, another polarity gene of *MARK* family, *MARK1*, has been recognized as susceptibility gene for autism,^49^ suggesting the involvement of polarity-related genes/pathways in ASD etiology.

The WNT signaling pathway mediates neurogenesis via several biological functions, including self-renewal, proliferation and differentiation.^39, 50–53^ For example, WNT3A and WNT7A are indispensable for maintaining self-renewal and stimulating the proliferation of neural stem cells,^50, 51^ and WNT3A also plays a critical role in neural fate commitment.^50^ The regulatory role of β-catenin signaling in the balance between cell proliferation and differentiation in the spinal cord was also reported.^54^ Protein□protein interaction (PPI) networks of ASD-associated genes have confirmed the convergent pathways underlying the development of ASD, including synaptic development, mitochondrial or metabolic processes, and WNT and MAPK signaling.^55–57^ Abnormal WNT signaling pathway activity has been reported in mouse models or patient-derived iPSCs carrying mutations in other high-confidence ASD genes, such as *SHANK3* and *TBR1*.^41, 58^. The transcriptomic profiling of our mutant iPSC-derived NPCs revealed reduced WNT/β-catenin signaling pathway. Moreover, decreased WNT3A, SOX1 and was seen in mutant neural rosettes. Our cellular phenotypes were also consistent with the developmental changes observed in neurons with *WNT3A/WNT7A* depletion.^50, 51, 53, 54, 59^ Furthermore, abnormal cellular phenotypes of mutant iPSC-derived NPCs were completely reversed *in vitro* by treatment with exogenous rhWNT3A. Our study, for the first time, links *MARK2* and the WNT signaling pathway, demonstrating that abnormal WNT/β-catenin signaling is the mechanism of *MARK2* loss.

To date, several molecules in the WNT/β-catenin signaling pathway, including *PTEN, ADNP, ARID1B, CHD8, ARID1B*,^5, 6, 57, 60^, the activator of β-catenin signaling *CTNNB1*^61^ and *WNT1*^62^, have been recognized as high-confidence ASD genes. The β-Catenin, as a highly conserved armadillo repeat protein family member, is encoded by *CTNNB1*, and Lof variants of *CTNNB1* cause a broad ASD phenotype.^61, 63^. To further confirm the involvement of the WNT/β-catenin signaling pathway in *MARK2*-related ASD, we compared the phenotypes of 120 individuals carrying germline likely pathogenic/pathogenic *CTNNB1* variants^61^ with those of our ASD individuals (**Supplementary Table 6**). The top rank phenotypes of *CTNNB1* variant are ID/DD (94.1%), motor delay (93.7%), delayed speech and language development/ASD (90.4%), and mild/severe eye abnormalities (87.5%). These highly penetrant phenotypes of *CTNNB1* variants are similar to that of *MARK2* variants, confirming the phenotypic similarity between *MARK2*-related and *CTNNB1*-related ASD.

Lithium, as a mood stabilizer, has been routinely prescribed to treat bipolar disorder for decades.^42, 43^ Previous *in vitro* and *in vivo* studies have proven that lithium directly activates the WNT/β-catenin signaling pathway.^41, 51, 64–67^ By activating the WNT/β-catenin signaling pathway, lithium promotes the proliferation of NPCs and improves behavioral performance in a mouse model of Down syndrome,^64^ and spatial memory impairment and neurodegeneration in a mouse model of Alzheimer’s disease.^67^ Lithium can also directly enhance the proliferation of hippocampal progenitors *in vitro* in a dose-dependent manner.^65^ The ability of lithium to rescue spine and synaptic defects has been reported in conditional *Tbr1*^+/-^ adult mice presenting ASD-like behaviors,^41^ and mice presenting impaired learning and memory.^66^ Siavash Fazel Darbandi generated ASD mice with conditional *Tbr1^layer^*^6^ knockout and found that both cellular and behavioral abnormalities, including immature dendritic spines reduced synaptic density, decreased social interactions between young mice, were rescued with LiCl treatment.^41^ In this study, we administered the clinically used dosage of lithium^31, 33^ to mutant iPSCs and *Mark2*^+/-^ (HET) mice. Abnormal cellular phenotypes in mutant NPCs, including alterations in the size of the neurosphere, rosette formation, the proliferation and differentiation of NPCs were reversed by lithium treatment. Considering that ASD is an early brain malformation, we fed a LiCl-treated diet to pregnant *Mark2*^+/-^ mice and studied the proliferation and differentiation of cortical neurons in *Mark2*^+/-^ mouse fetuses. Abnormal cortex layer formation was also reversed. In the future, the ASD-related behaviors of *Mark2*^+/-^ offspring mice fed a LiCl-containing diet during the prenatal/postnal period will be studied to confirm the therapeutic potential of LiCl for ASD.

### Conclusion

Our studies deciphered the phenotypic and variant spectrum of *MARK2*-related ASD. Using human iPSC-derived NPCs *in vitro* and model mice *in vivo*, we elucidated the cellular phenotypes and molecular mechanism of *MARK2* loss during early neural development which are associated with down-regulation of the WNT/β-catenin signaling pathway. Moreover, we observed that lithium can reverse cellular phenotype in mutant iPSC-NPCs and abnormal cortical partition development in knockout mice via activating the WNT/β-catenin signaling pathway, providing a potential treatment for *MARK2*-related ASD patients.

## Acknowledgements

We thank all the families who participated in this study. We also thank James F. Gusella for his valuable comments and suggestions.

## Author contribution

Maolei Gong, Chang-Mei Liu and Xiaoli Chen were responsible for conception and design. Maolei Gong, Xiaoli Chen, Changmei Liu, Jiayi Li, Yijun Liu contributed to manuscript writing, and Matheus Vernet Machado Bressan Wilke and Michael T Zimmermann help in manuscript editing. Xiaoli Chen, Zailong Qin, Vernet Machado Bressan Wilke, Matheus, Jiayi Li, and Haorang Liu managed the analysis and interpretation of all clinical/genetic data. Jiayi Li and Haoran Liu performed the WB and qPCR experiments for PBMCs, in vitro 293HEK/Hela transfection assay for the spicing and missense variants. Maolei Gong, Yijun Liu, Jiayi Li, Qian Li and Chen Liang performed the iPSC-related experiments. Maolei Gong, Yijun Liu and Haoran Liu conducted the animal experiments. Michael T Zimmermann, Neshatul Haque, Nikita R. Dsouza, and Raul Urrutia performed structural modeling. Hongzhen Du helped in iPSC culture and neural differentiation, drug treatment for iPSCs and HET mice. Maolei Gong analyzed and visualized the RNA-Seq data. All authors took part in data collection and analysis and approved the final manuscript.

The following partners conducted inviduals recruitment and clinical/genetic information and photo collection (listed in alphabetical order): Ana S.A. Cohen, Benjamin Cogne, Bernt Popp, Bonnie R. Sullivan, Boris Keren, Carrie Lahner, Changhong Ren, Christiane Zweier, Christine Coubes, Claudine Rieubland, Daniel C. Koboldt, Dominique Braun, Ellen van Binsbergen, Eric W Klee, Frederic Tran Mau-Them, Hana Safraou, Henry Oppermann, Hua Xie, Jacques Michaud, Jianbo Zhao, Joel A Morales-Rosado, Julie Gauthier, Juliette Piard, Koen L van Gassen, Konrad Platzer, Laïla El Khattabi, Lucie Evenepoel, Marie-José H. van den Boogaard, Marjolaine Willems, Matheus Vernet Machado Bressan Wilke, Matthew J. Ferber, Micheil Innes, Michelle D. Amaral, Mignot Cyril, Nicole J Boczek, Odent Sylvie, Pia Zacher, Pierre Blanc, Richard H. van Jaarsveld, Scott Douglas McLean, Sherr Elliott, Susan M. Hiatt, Susan S. Hughes, Tianyun Wang, Valerie Waddell, Weixing Feng, Wen-Hann Tan, Whitley V. Kelley, Xiaoyan Wang, Yazhen Yu, Yiping Shen, Yiyan Ruan, Zailong Qin.

## Supplementary material

Supplementary material is available online.

