## supplementary materials for "*MARK2* variants cause autism spectrum disorder *via* the downregulation of WNT/β-catenin signaling pathway"

**Subtitle: Deciphering clinical features and mechanism of *MARK2*-related ASD**

Maolei Gong [1,2,3,57,†], Jiayi Li [4,5,†], Yijun Liu [1,2,3,6,†], Matheus Vernet Machado Bressan Wilke [7,†], Qian Li [1,2,3,6], Haoran Liu [4], Chen Liang [4], Joel A Morales-Rosado [8], Ana S.A. Cohen [9,10], Susan S. Hughes [10,11], Bonnie R. Sullivan [10,11], Valerie Waddell [12], Marie-José H. van den Boogaard [13], Richard H. van Jaarsveld [13], Ellen van Binsbergen [13], Koen L van Gassen [13], Tianyun Wang [14,15], Susan M. Hiatt [16], Michelle D. Amaral [16], Whitley V. Kelley [16], Jianbo Zhao [17], Weixing Feng [17], Changhong Ren [17], Yazhen Yu [18], Nicole J Boczek [19], Matthew J. Ferber [19], Carrie Lahner [19], Sherr Elliott [20], Yiyan Ruan [21], Mignot Cyril [22], Boris Keren [22], Hua Xie [4], Xiaoyan Wang [23], Bernt Popp [24,25], Christiane Zweier [26,27], Juliette Piard [28,29], Christine Coubes [30], Frederic Tran Mau-Them [31,32], Hana Safraou [31,32], Micheil Innes [33], Julie Gauthier [34,35], Jacques Michaud [35,36], Daniel C. Koboldt [37], Odent Sylvie [38,39], Marjolaine Willems [40], Wen-Hann Tan [41], Benjamin Cogne [42], Claudine Rieubland [26], Dominique Braun [26], Scott Douglas McLean [43,44], Konrad Platzer [45], Pia Zacher [46], Henry Oppermann [45], Lucie Evenepoel [47], Pierre Blanc [48], Laïla El Khattabi [49], Neshatul Haque [50], Nikita R. Dsouza [50], Michael T. Zimmermann [50,51,52], Raul Urrutia [53], Eric W Klee [7,54], Yiping Shen [41,55], Hongzhen Du [1,2,3], Zailong Qin [56*], Chang-Mei Liu [1,2,3,6*] and Xiaoli Chen [4, 5*]

**†** Maolei Gong, Jiayi Li, Yijun Liu and Matheus Vernet Machado Bressan Wilk contributed equally to this work.

1 State Key Laboratory of Stem Cell and Reproductive Biology, Institute of Zoology, Chinese Academy of Sciences, Beijing, China.

2 Beijing Institute for Stem Cell and Regenerative Medicine, Beijing, China.

3 Institute for Stem Cell and Regeneration, Chinese Academy of Sciences, Beijing, China.

4 Department of Medical Genetics, Capital Institute of Pediatrics, Beijing, China.

5 Chinese Academy of Medical Sciences & Peking Union Medical College, Beijing, China.

6 Savaid Medical School, University of Chinese Academy of Sciences, Beijing, China.

7 Department of Clinical Genomics, Mayo Clinic, Rochester, MN, USA.

8 Department of Pathology, Microbiology & Immunology, Vanderbilt University Medical Center, Nashville, TN 37232, USA.

9 Department of Pathology and Laboratory Medicine, Genomic Medicine Center, Children's Mercy-Kansas City, Kansas City, MO, USA.

10 The University of Missouri-Kansas City, School of Medicine, Kansas City, MO, USA.

11 Division of Clinical Genetics, Children's Mercy Kansas City, Kansas City, MO, USA.

12 Department of Neurology, Children's Mercy Kansas City, Kansas City, MO, USA.

13 Department of Genetics, University Medical Center Utrecht, Heidelberglaan 100, 3584 CX Utrecht, the Netherlands.

14 Department of Medical Genetics, Center for Medical Genetics, School of Basic Medical Sciences, Autism Research Center, Peking University Health Science Center, Beijing, China. 15 Neuroscience Research Institute, Peking University; Key Laboratory for Neuroscience, Ministry of Education of China & National Health Commission of China, Beijing, China.

16 HudsonAlpha Institute for Biotechnology, Huntsville, AL, USA.

17 Department of Neurology Beijing Children's Hospital, Capital Medical University, Beijing, China.

18 Department of Pediatrics, Beijing Tiantan Hospital affiliated with Capital University of Medical Sciences, Beijing, China.

19 Department of Laboratory Medicine and Pathology, Genomics Laboratory, Mayo Clinic, Rochester, MN, USA.

20 Departments of Neurology and Pediatrics, Institute of Human Genetics and Weill Institute for Neurosciences, University of California, San Francisco, California, USA.

21 Guangxi Clinical Research Center for Pediatric Diseases, The Maternal and Child Health Care Hospital of Guangxi Zhuang Autonomous Region,

22 Department of Medical Genetics, Pitié-Salpêtrière Hospital, AP-HP, Sorbonne Université, Paris, France.

23 Department of Children's Nutrition Research Center, Affiliated Children’s Hospital of Capital Institute of Pediatrics, Beijing, China.

24 Institute of Human Genetics, University of Leipzig Hospitals and Clinics, Leipzig, Germany.

25 Berlin Institute of Health at Charité-Universitäts medizin Berlin, Center of Functional Genomics, Hessische Straße 4A, Berlin, Germany.

26 Department of Human Genetics, Inselspital, Bern University Hospital, University of Bern, Bern, Switzerland.

27 Institute of Human Genetics, University Hospital Erlangen, Friedrich-Alexander Universität Erlangen-Nürnberg, Erlangen, Germany.

28 Centre de Génétique Humaine, Centre Hospitalier Régional Universitaire, Université de Franche-Comté, Besançon, France.

29 UMR 1231 GAD, Inserm, Université de Bourgogne Franche Comté, Dijon, France.

30 Département de Génétique Médicale, Maladies Rares et Médecine Personnalisée Hôpital Arnaud de Villeneuve 34295 MONTPELLIER Cedex 5, 21000 Dijon, France.

31 UF6254 Innovation en Diagnostic Genomique des Maladies Rares, 21070 Dijon, France.

32 Inserm UMR1231 GAD, F-21000, Dijon, France.

33 Department of Medical Genetics and Pediatrics and Alberta Children's Hospital Research Institute, Cumming School of Medicine, University of Calgary.

34 Molecular Diagnostic Laboratory, Centre Hospitalier Universitaire Sainte-Justine, Montréal, QC, Canada.

35 Department of Pediatrics, Université de Montréal, Montréal, QC, Canada.

36 CHU Sainte-Justine Research Center, Montreal, Quebec, Canada.

37 The Institute for Genomic Medicine, Nationwide Children's Hospital.

38 Service de Génétique clinique, CHU Rennes, ERN ITHACA, France.

39 Univ Rennes, CNRS, INSERM, IGDR (Institut de Génétique et développement de Rennes), UMR 6290, ERL U1305, Rennes, France.

40 Medical Genetic Department for Rare Diseases and Personalized Medicine, Reference Center AD SOOR, AnDDI-RARE, Inserm U1298, INM, Montpellier University, Centre Hospitalier Universitaire de Montpellier, 34090 Montpellier, France.

41 Division of Genetics and Genomics, Boston Children's Hospital, Harvard Medical School.

42 Nantes Université, CHU Nantes, Service de Génétique Médicale, Nantes, France; Nantes Université, CHU Nantes, CNRS, INSERM, l'institut du thorax, Nantes, France.

43 Division of Clinical Genetics, The Children's Hospital of San Antonio, San Antonio, TX 78207, USA.

44 Department of Molecular and Human Genetics, Baylor College of Medicine, Houston TX 77030, USA.

45 Institute of Human Genetics, University of Leipzig Medical Center, Leipzig, Germany.

46 Epilepsy Center Kleinwachau, 01454 Dresden-Radeberg, Germany.

47 Centre de Génétique Humaine, Cliniques Universitaires Saint-Luc, Université Catholique de Louvain, Avenue Hippocrate 10-1200, Brussels, Belgium.

48 Sorbonne Université, Department of Medical Genetics, APHP, Pitié-Salpêtrière hospital, Paris Brain Institute-ICM, Laboratoire SeqOIA-PFMG2025, 75014 Paris, France.

49 Department of Medical Genetics, APHP, Armand Trousseau and Pitié-Salpêtrière hospitals, Brain Development team, Paris Brain Institute-ICM, Sorbonne Université, 75013, Paris, France; Laboratoire SeqOIA-PFMG2025, 75014 Paris, France.

50 Bioinformatics Research and Development Laboratory, Linda T. and John A. Mellowes Center for Genomic Sciences and Precision Medicine, Medical College of Wisconsin, Milwaukee, WI 53226, USA.

51 Department of Biochemistry, Medical College of Wisconsin, Milwaukee, WI, USA.

52 Clinical and Translational Sciences Institute, Medical College of Wisconsin, Milwaukee, WI, USA.

53 Department of Surgery, Medical College of Wisconsin, Milwaukee, WI, USA.

54 Department of Quantitative Health Sciences, Mayo Clinic, Rochester, MN, USA.

55 Department of Neurology, Harvard Medical School, Boston, MA, USA.

56 Genetic and Metabolic Central Laboratory, Birth Defect Prevention Research Institute, Maternal and Child Health Hospital of Guangxi Zhuang Autonomous Region, Nanning, China.

57 Strategic Support Force Medical Center, Beijing 100024, China.

Correspondence to:

Zailong Qin, Genetic and Metabolic Central Laboratory, Birth Defect Prevention Research Institute, Maternal and Child Health Hospital of Guangxi Zhuang Autonomous Region, Nanning 530002, China. 86-771-3152428,.

Chang-Mei Liu, Institute of Zoology, Chinese Academy of Sciences, 1 Beichen West Road, Chaoyang District, Beijing,100101, China, 86-10-82619690,.

Xiaoli Chen, Capital Institute of Pediatrics, No. 2, Yabao Road, Chaoyang District, Beijing, 100020, P.R.China, 86-10-85695525,.

The authors have declared that no conflict of interest exists.

### METHODS

#### Behavioral and memory tests

All mice used for the behavioral tests were male mice aged 8-12 weeks (n≥8 per group), and all tests were performed between 09:00 and 17:00. Videos of the behavioral tests were analyzed by EthoVision XT 14 (Noldus).

#### Open field test

The open field test was conducted in a 50 x 50 x 50 cm box. A test subject was placed in the center of the box, and its behavior was video recorded for 5 min by a camera positioned directly above the box. Total distance and speed were quantified during video recording. A 30 cm square was delineated as the center zone.

#### Elevated plus maze test

Each mouse was placed in the central area of the elevated plus maze facing one of the open arms. The mice were allowed to explore the maze for 5 min, and the time spent in the open arm was calculated with EthoVision XT 14.

#### Three-chamber test

Two weeks before testing, subject animals and stimulus animals (female C57BL/6 mice aged 3-4 months) were housed alone in individual clean cages in the testing room. The test was performed in a novel, clean box (72 cm length x 72 cm width x 36 cm height) during the light phase. A subject mouse was placed in the empty apparatus and allowed to habituate to the three chambers, which contained two transparent bottles with holes (9 cm in diameter, 12 cm high), for 15 min. After habituation, one female stimulus mouse was placed in one of the transparent bottles in the right chamber, and the subject mouse was placed in the middle chamber and allowed to explore for 15 min. The subject mouse was then returned to its home cage. Then, another unknown female mouse was placed in the bottle in the other chamber (left) of the test box; the subject mouse was placed back in the test box equidistant from and facing the familiar and novel female mice. Interactions between the subject mouse with the familiar and novel female mice were videotaped for 15 min. Sniffing times were recorded and analyzed.

#### Novel object recognition test

A mouse was placed as in an open field test arena facing a wall and allowed to freely explore for 5 min. After a short rest in its home cage, the mouse was placed in the box again facing two identical objects (5 cm away from the walls) and allowed to explore for another 10 min exploration (T1). After a 60-min rest period, the mouse was placed in the box again facing two objects (a novel one and one that was present in T1) and allowed to explore for another 10 min (T2). The recognition index was calculated and analyzed.

**Marble-burying test**

Mice were individually placed in Plexiglas cages containing 5-cm-deep fresh bedding, and then 20 black glass marbles (15 mm diameter) were gently placed in a 4 x 5 arrangement at equal distances. Testing was conducted for 30 min. After the test period, buried marbles were counted. Marbles were considered buried if at least one half was covered with bedding.

**Grooming test**

The grooming task consisted of 15 min of habituation followed immediately measurement of grooming behavior for 15 min. Mice were individually placed in novel Plexiglas cages (45 cm x 22 cm). The time spent grooming the genitals, tail, paw, leg, body and head was recorded in seconds.

**Y-maze test**

Mice were placed in one arm of the Y-maze apparatus (20 cm high, 50 cm long, and 10 cm wide at the bottom) and allowed to explore freely for 10 min. A correct spontaneous alternation was defined as the successive entry of a mouse into the three arms in overlapping triplet sets. The spontaneous alternation percentage (%) was calculated as the number of successive triplet sets (consecutive entries into three different arms)/total number of arm entries minus 2) x100.

**Barnes maze test**

The apparatus was a rotatable gray acrylic disc (1.22 m in diameter) elevated 0.58 m above the floor with 20 holes (5 cm diameter, 2 cm away from the edge) equally spaced along the perimeter. The apparatus was brightly lit (600 lux). Only one hole in the maze top led to a removable hiding box, which was situated directly below the escape hole. On the first day, a mouse was placed in a transparent cylindrical start chamber (10.5 cm), and 30 s after the onset of a buzzer sound (85 dB), the chamber was lifted, and the mouse was allowed to freely explore the maze. The trial ended when the mouse entered the hiding box or after 3 min had elapsed. Mice that did not find the hiding box by the end of the 3-min period were gently guided to the escape hole by the investigator. Immediately after the mouse entered the hiding box, the buzzer sound was turned off, and the mouse was allowed to stay in the hiding box for 1 min. On the second day, the mouse was placed in a black square-shaped start chamber (10.5 cm), and 15 s after the onset of a buzzer sound (85 dB), the chamber was lifted, and the mouse was allowed to freely explore the maze. The trial ended when the mouse entered the hiding box or after 2 min had elapsed. Immediately after the mouse entered the hiding box, the buzzer sound was turned off, and the mouse was allowed to stay in the hiding box for 1 min. The experiment was repeated three times for each mouse. The procedure performed on the second day was conducted twice on the third day. On the fifth day, the subjects were placed in a black square-shaped start chamber (10.5 cm), and 15 s after the onset of a buzzer sound (85 dB), the chamber was lifted, and the mouse was allowed to freely explore the maze. The trial ended when the mouse entered the hiding box or after 2 min had elapsed. Immediately after the mouse entered the hiding box, the buzzer sound was turned off. The movements of the animals in the maze were digitally recorded.

**Morris water maze test**

A 120 cm diameter, 45 cm deep Morris water maze was filled with water, which was made with nontoxic white paint, to a depth of 25 cm. An escape platform (diameter 13 cm) was hidden 1 cm beneath the surface of the water in the center of one of the quadrants of the water tank. Four extra-maze cues, i.e., different shapes, were placed at equal distances on the wall surrounding the water tank. The water temperature was adjusted to 21±1°C. The mice were trained to find the escape platform in four trials per day for 6 consecutive days. In each trial, a mouse was placed in a randomly chosen quadrant and allowed to swim for up to 1 min to find and climb on the platform. If it failed to find the platform within that time, it was guided to the escape platform and kept there for 15 s. A probe test was conducted 24 h after completion of training. In the probe test, the platform was removed from the pool, and behavior was recorded for 60 s. Latency to reach the platform and time spent in the platform quadrant were recorded.

A

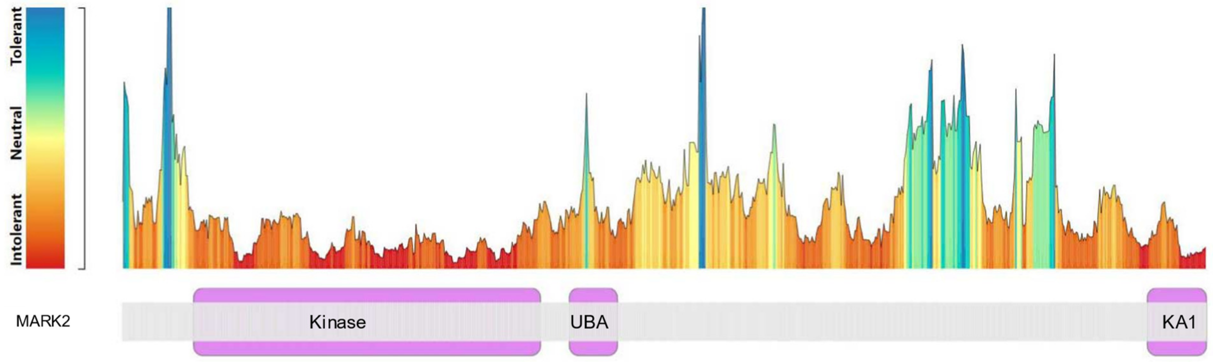


**Figure S1. The missense variant tolerance landscape**

Protein of MAKR2 (GENCODE: ENST00000402010.2, RefSeq: NM_001039469.2, UniProt: Q7KZI7) was used. The ratios of missense over synonymous variant were calculated to indication of regions that are intolerant to missense variation. The kinase and KA1 domain (purple parts) clearly show as intolerant compared with other parts.

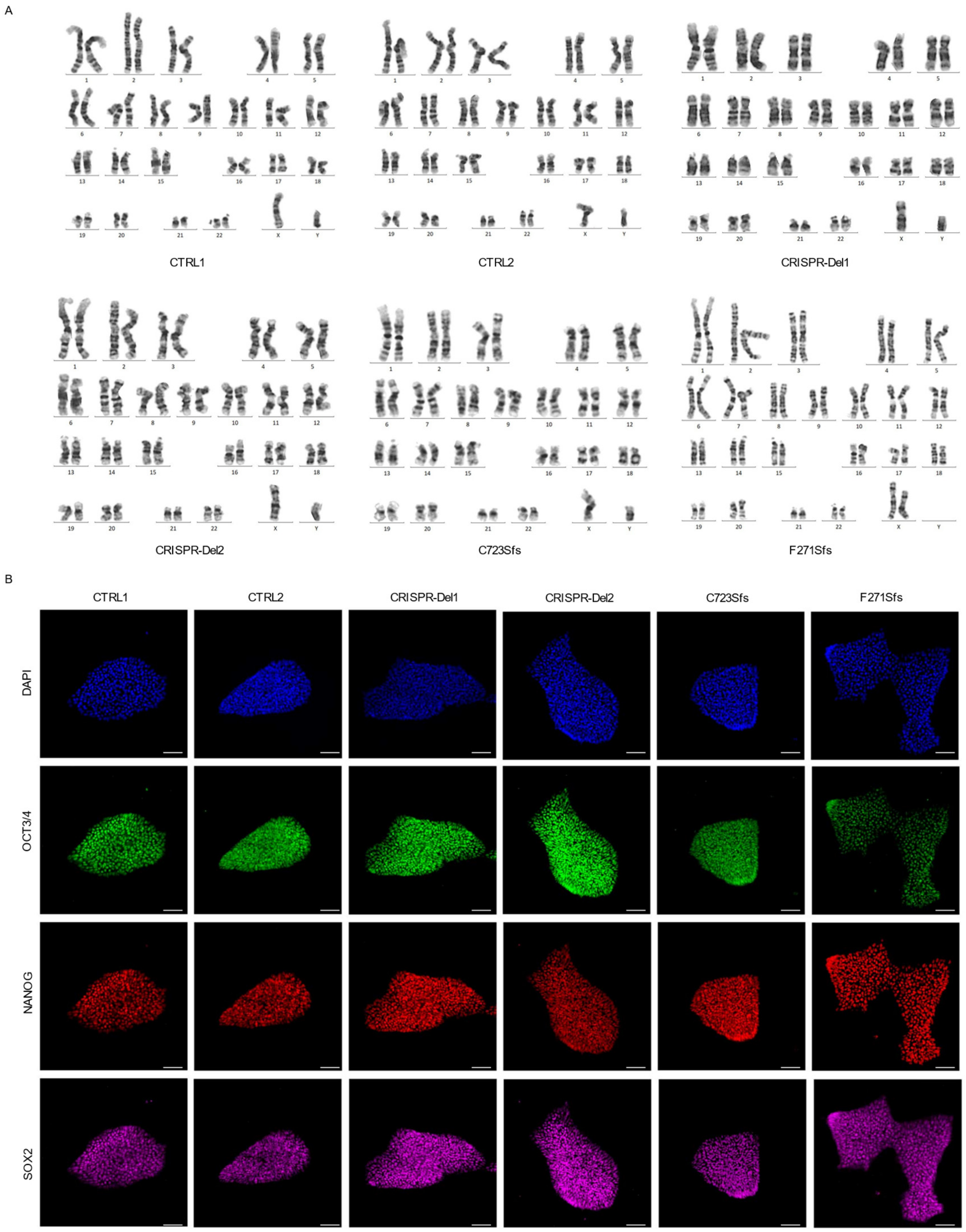


**Figure S2. Representative validation images of iPSCs**

**A**, Representative karyotype image of six iPSCs with different genotype. CTRL1 and CTRL2: two independent healthy adults without *MARK2* variant; C723Sfs and F271Sfs: two affected individuals with *MARK2* variant; CRISPR-Del1 and CRISPR-Del2: two isogenic *MARK2* deletion produced by the CRISPR/Cas9 editing technology. **B**, Representative validation images of six iPSCs using several markers, OCT 3/4 (green), NANOG (red), and SOX2 (purple). Nuclei were stained with DAPI (blue). scale bar = 50 μm.

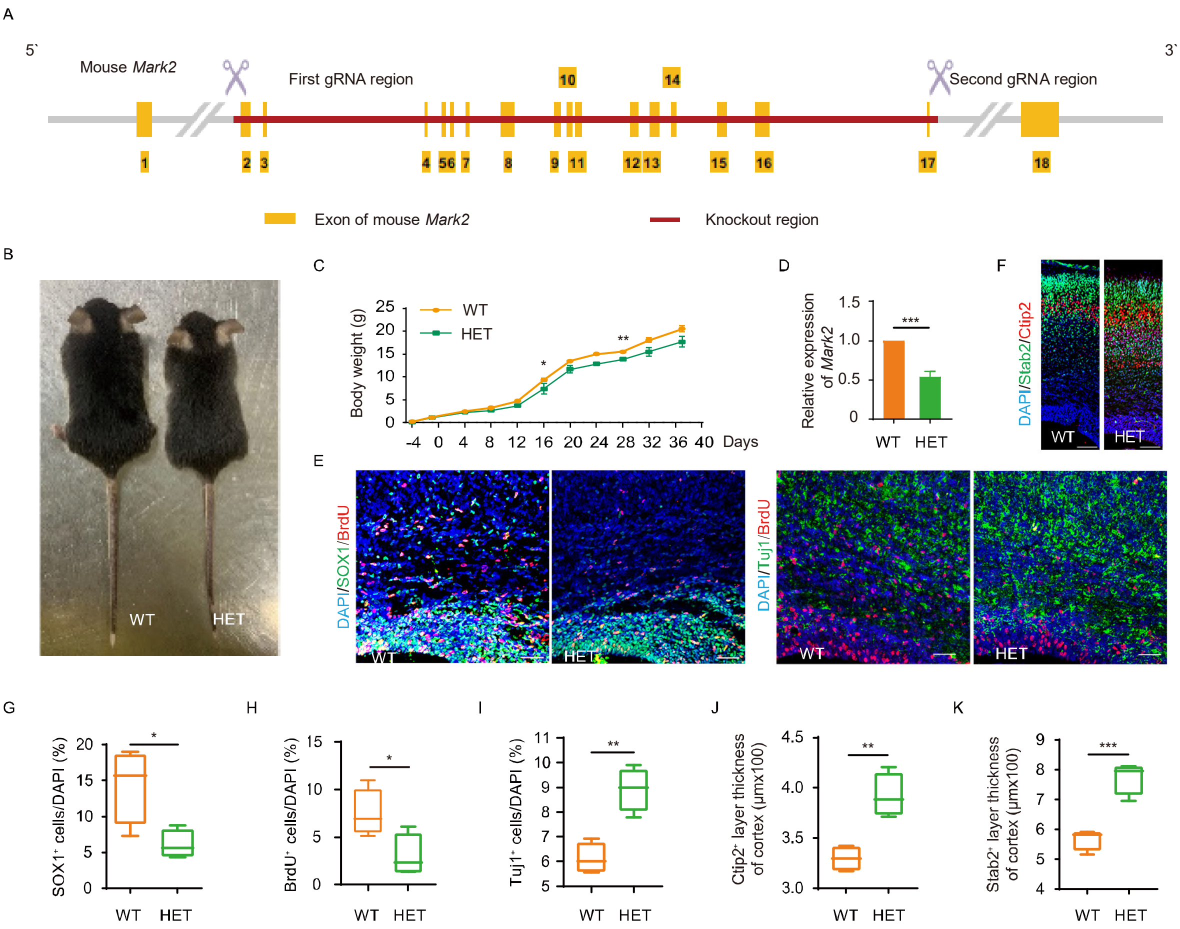


**Figure S3. *Mark2* loss in mice affects the proliferation and differentiation of NPCs *in vitro.***

**A,** Schematic of *Mark2* knockout mice. **B-C**, Body size (**B**) and growth curve (**C**) of *Mark2*$^{+/+}$ (WT) mice and *Mark2*$^{+/-}$ (HET) mice in 6 weeks. **D,** Expression of *Mark2* of HET and WT mice. Total RNAs were isolated from the cortex in E18.5 mice, and GAPDH was used as the internal parameter. **E-F**, Representative images of immunofluorescence staining for BrdU (red) and SOX1 (green), BrdU (red) and TUJ1 (green), Ctip2 (red) and Stab2 (green) in the cortical region above the subventricular zone (SVZ) in E18.5 mice. **G-K**, Quantification analysis of SOX1$^{+}$ cells (**G**), BrdU$^{+}$ cells (**H**), TUJ1$^{+}$ cells (**I**), Ctip2$^{+}$ cells (**J**), and Stab2$^{+}$ cells (**K**). WT=5, HET=4. The data are presented as the mean±s.e.m. of at least three independent experiments and were analyzed by Student's t test; **p*<0.05, ***p*<0.01 and ****p*<0.001. scale bar = 50 μm.

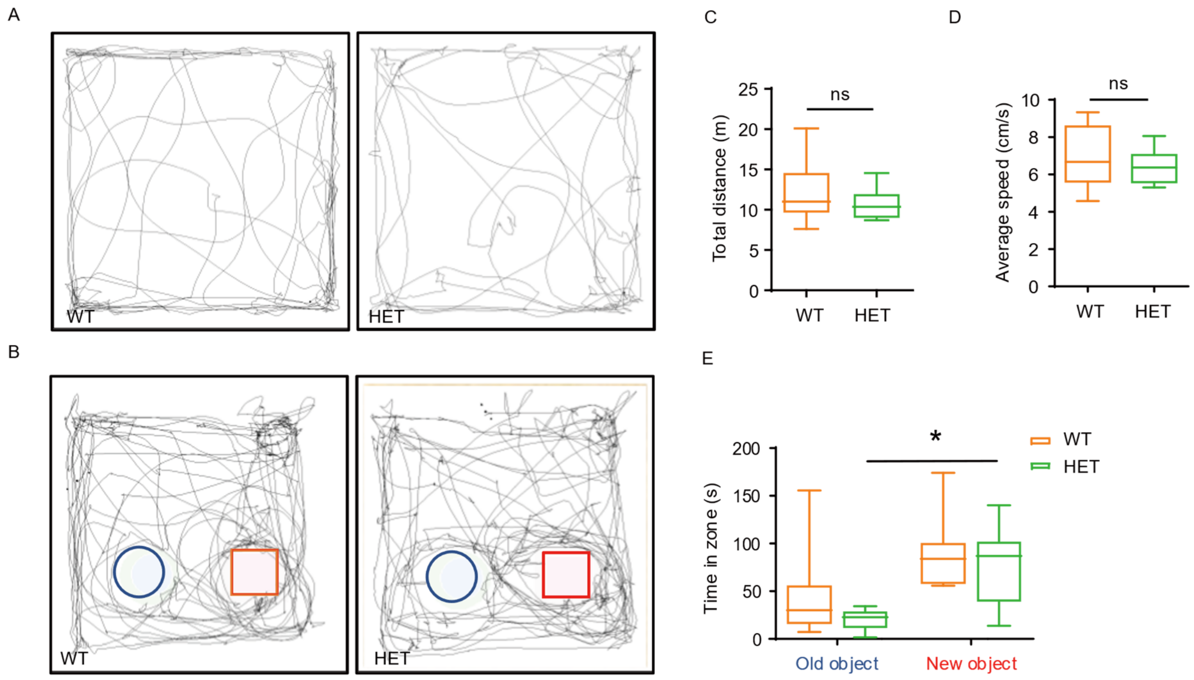


**Figure S4. Open field and new object recognition tests in mice with *Mark2* loss**

**A-B**, Trajectories of WT and HET mice in the open field test (**A**) and new object recognition test. (**B**). **C-D**, Quantification analysis of total distance (**C**) and average speed (**D**) between two genotypes in the open field test. **E**, Quantification analysis of time in zone in the new object recognition test ($n$=11). Time spend in contact with the object was used to represent for the new object recognition. The blue circle was used to point the location of the old object and red square was used to point the location of the new object. WT=11, HET=11. The data are expressed as the mean±s.e.m. of at least three independent experiments and were analyzed by Student's t test; *$p$<0.05. ns: not significant.

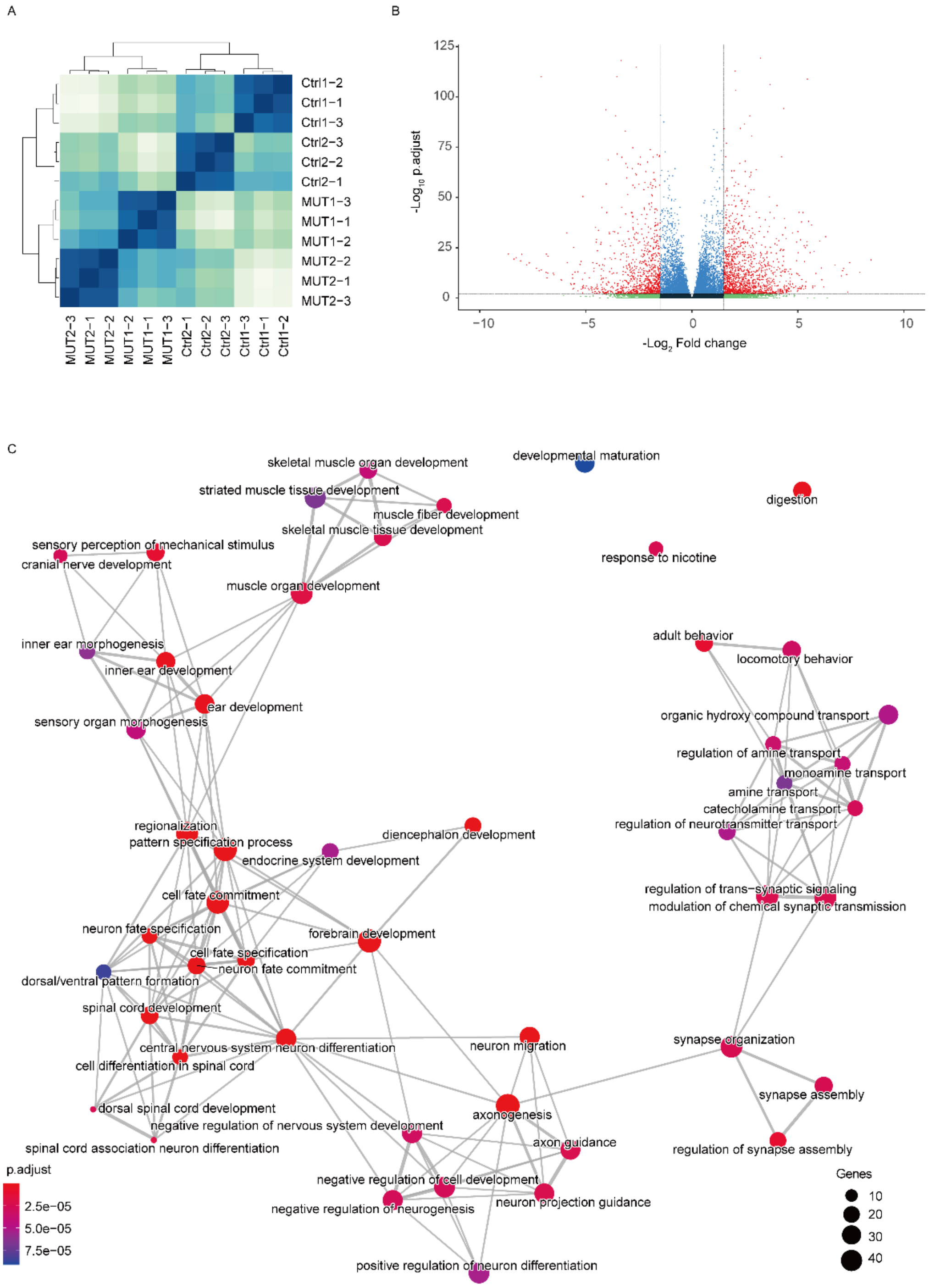


**Figure S5. RNA-Seq and Gene ontology (GO) analyses of iPSC-derived NPCs.**

**A**, Correlation heatmap of RNA-Seq of control (CTRL) and mutant iPSC (C723Sfs)-derived NPCs (N=3 experiment). **B**, Example volcano plot. Points on top-right and top-left corners are considered the most promising genes (*P*<0.05, $log_2$Foldchange>1.5). **C**, Association network diagram between feature sets through GO analysis. All those feature sets of GO were enriched through downregulated genes ($Log_2$FoldChange<-1.5, *p* value>0.05) in mutant neurospheres.

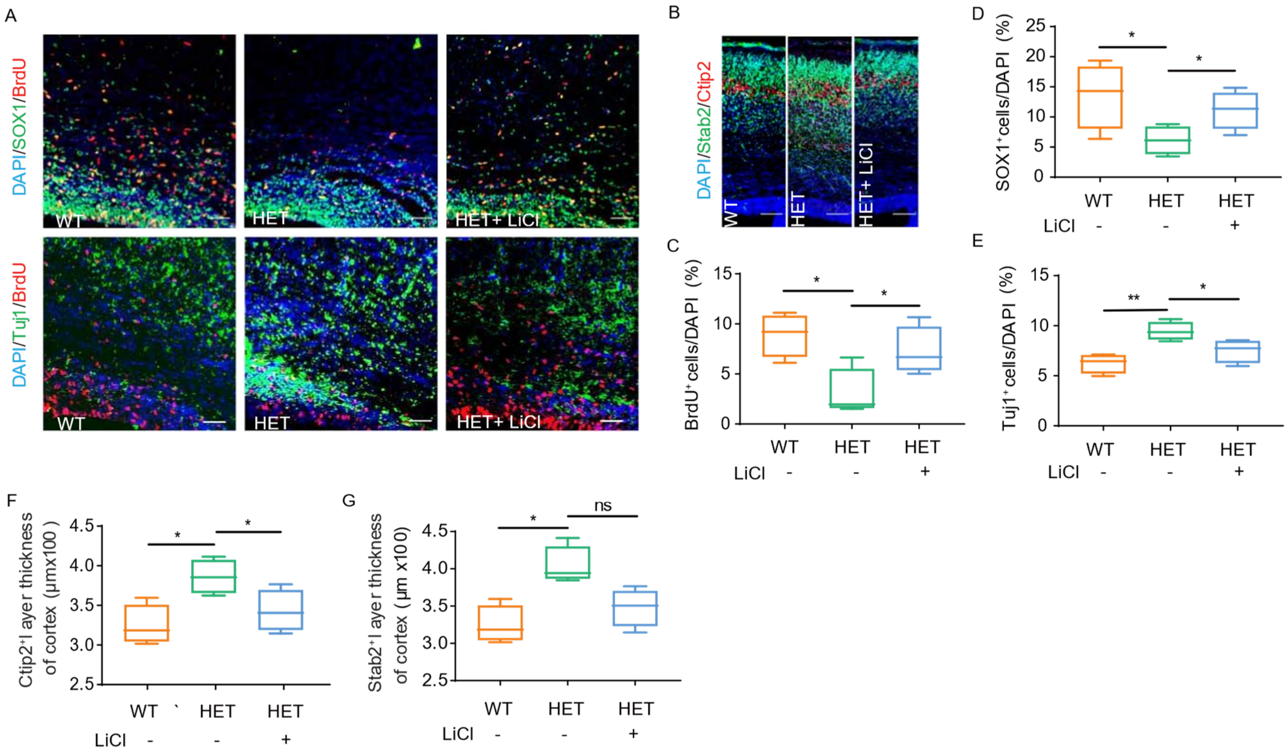


**Figure S6. Abnormal cortical development in *Mark* $^{+/-}$ mice is rescued by LiCl**

**A-B,** Representative images of immunofluorescence staining for BrdU/SOX1, BrdU/TUJ1 (**A**), and Ctip2/Stab2 (**B**) in the mouse embryonic cortex (E18.5) from three groups: *Mark2*$^{+/+}$ mice (WT=4), untreated *Mark2*$^{+/-}$ mice (HET=4) and LiCl-treated *Mark2*$^{+/-}$ mice (HET+LiCl=4). **D-G,** Quantification analysis of the data **in A-B,** including the numbers of SOX1$^{+}$ cells (**D**), BrdU$^{+}$ cells (**C**), and TUJ1$^{+}$ cells (**E**), Ctip2$^{+}$ layer (**F**) and Stab2$^{+}$ layer thickness (**G**). The data are expressed as the mean±s.e.m of at least three independent experiments and were analyzed by Student's t test; *$p<0.05$, and **$p<0.01$. ns: not significant. scale bar = 50 μm.
